# Machine Learning Approaches to Predict Alcohol Consumption from Biomarkers in the UK Biobank

**DOI:** 10.1101/2024.12.22.24319486

**Authors:** Mohammed F. Hassan, Amanda Elswick Gentry, Elizabeth C. Prom-Wormley, Roseann E. Peterson, Bradley T. Webb

## Abstract

**Background:** Measuring and estimating alcohol consumption (AC) is important for individual health, public health, and Societal benefits. While self-report and diagnostic interviews are commonly used, incorporating biological-based indices can offer a complementary approach.

**Methods:** We evaluate machine learning (ML) based predictions of AC using blood and urine-derived biomarkers. This research has been conducted using the UK Biobank (UKB) Resource. In addition to the prediction of the number of alcoholic Drinks Per Week (DPW), four other related phenotypes were predicted for performance comparison. Five ML models were assessed including LASSO, Ridge regression, Gradient Boosting Machines (GBM), Model Boosting (MBOOST), and Extreme Gradient Boosting (XGBOOST).

**Results:** All five ML methods achieved moderate prediction of DPW (r^2^=0.304-0.356) with biomarkers significantly increasing prediction above using only known covariates and liver enzymes (r^2^=0.105). XGBOOST achieved the best prediction performance (r^2^=0.356, MAE=5.214) at the expense of increasing model complexity and training resources compared to other ML methods. All ML models were able to accurately predict if subjects were heavy drinkers (DPW>8 for women and DPW>15 for men) and produced explainable models that highlighted the role of biomarkers in predicting DPW. While phenotype correlations were similar across methods, XGBOOST produced similar heritability estimates for observed (h^2^=0.064) and predicted (h^2^=0.077) DPW. The estimated genetic correlation between observed and predicted DPW was 0.877.

**Conclusions:** Predicting AC from ML-based biological measures provides an opportunity to identify individuals at increased risk of heavy AC, thereby offering complementary avenue for risk assessment beyond self-report, screening instruments, or structured interviews, which have some known biases. In addition, explainable AI tools identified a constellation of biomarkers associated with AC.

## Introduction

The ability to measure alcohol consumption (AC) across multiple contexts is important, including research, individual health [1], public health [2], and social policy [3]. Alcohol consumption also has a complex relationship with mental health outcomes and psychiatric disorders [4]. While the impact of excessive consumption and alcohol use disorder (AUD) on health is well documented and causally linked to liver damage, cardiovascular issues, pancreatitis, and more [5], the role of lower levels of consumption contributing to adverse outcomes is still debated. Recently, moderate drinking has not been shown to be protective [6] and the WHO concluded there is no safe level of alcohol consumption. Given this context, the accurate measurement or estimation of AC across the full range of consumption is increasingly important to researchers, clinicians, and public health officials. Identifying associations and patterns of alcohol consumption, particularly on the higher end, is crucial for designing targeted interventions and prevention programs that allow for better allocation of resources for treatment, education, and support services [7].

### Structured interviews and self-report

While surveys and structured interviews are considered a standard method for assessing alcohol consumption, alcohol problems, and diagnosing Alcohol Use Disorder (AUD) [8] [9], they have limitations including relying on the accuracy of self-report. Responses can be influenced by factors such as memory biases, social desirability, or reluctance to disclose certain information leading to potential underreporting or misdiagnosis, among others. Biological-based indices or biomarkers may offer complementary information to self-report based measures and diagnoses since biomarkers can reflect physiological or neurobiological changes associated with specific conditions [10].

### Utility and need for biomarker-based estimations of alcohol consumption

There are a variety of avenues to assess alcohol consumption without biomarkers. However, the estimates of average consumption as well as the prevalence of heavy consumption, binge drinking, and AUD can vary greatly depending on the method and dataset. The National Epidemiologic Survey on Alcohol & Related Conditions (NESARC-III) is a large representative sample of US adults. NESARC conducted face-to-face interviews and deep assessments of alcohol and drug use, related risk factors, and associated physical and mental disabilities in 36,309 participants [11]. While NESARC represents a high-quality assessment, reported drinks per day can differ between last year versus lifetime maximum within the same individual.

Studies using structured interviews, such as NESARC-III, show lifetime AUD prevalences of ∼29% [12]. The recently reported prevalence of AUD in All of Us is 1.88% when using an EHR-based ascertainment approach [13] indicating that standard surveying incorporated into routine medical care may not accurately reflect alcohol-related behaviors. This discrepancy may significantly impact alcohol-related research since misclassification reduces the statistical power of both epidemiological and genetic discovery of risk. Since structured interviews may not be feasible in large biobanks, there is a need to assess if available objective biological measures can be used to augment EHR and self-report variables describing alcohol-related outcomes.

### Known biomarkers of alcohol consumption

While ethanol can be measured directly in serum, plasma, blood, and urine, this measurement only reflects recent acute ethanol consumption and is not an index of regular or heavy consumption. In contrast, other blood-based measures are correlated with frequent and persistent heavy consumption [14], including ethyl glucuronide, ethyl sulfate, phosphatidylethanol (PEth), and liver enzymes [15]. Many other biomarkers have been proposed and evaluated [16], however, there is no consensus regarding the best set of markers, and there are no well-established algorithms to predict alcohol consumption, problems, or use disorder using objective biological measures. However, established and commonly assessed alcohol-related biomarkers such as aspartate aminotransferase (AST), alanine aminotransferase (ALT), and gamma-glutamyl transferase (GGT) may still be useful [17]. In patients being treated for AUD, these measures provide converging evidence that self-reported alcohol consumption is consistently under-reported [18]. To our knowledge, there are no validated biomarkers that can quantitatively estimate lower levels of AC, which may still impact health outcomes. This highlights the need for reliable tools for the estimation of alcohol consumption across its full range.

### Introduction Summary

We have previously demonstrated that the Alcohol Use Disorders Identification Test (AUDIT) can be predicted using a large number of variables with complex patterns of missingness [19]. This research sought to determine if objective biological measures beyond self-reporting can predict alcohol consumption using machine learning (ML) algorithms. A successful prediction model would facilitate the development of a complementary screening tool for alcohol consumption where self-report may not be available or reliable. Specifically, we applied a series of ML algorithms to predict alcoholic drinks per week (DPW) using panels of urine and blood chemistry markers (hereafter referred to as “biomarkers”) in a sample from the UK Biobank (UKB.) These predictions are validated using genetic correlation (r_G_) estimates between observed, measured phenotypes, and ML-predicted phenotypes.

Predictive models for AC offer several advantages across various contexts, including imputing missing alcohol consumption variables and predicting alcohol consumption from biological markers in cases where AC may not be directly assessed in the clinical encounter and/or where true consumption habits may be underreported. Where appropriate, such predictions can increase sample sizes and improve the power of epidemiological and genetic association discovery.

Furthermore, ML models facilitate the creation of interpretable models, enabling the identification and interpretation of biomarkers and other variables linked to AC, and could enable further investigations.

## Methods

### Sample

This research has been conducted using the UK Biobank (UKB) Resource (application number 30782). For phenotype predictions, the input variables included 249 Nuclear Magnetic Resonance (NMR) biomarkers from plasma, 30 blood biochemistry measures, 31 blood count measures, 25 infectious disease blood measures, and 3 urine assay measures for a total of 338 initial predictors (Supplementary Tables T1 and T2). The NMR biomarkers consist of lipoprotein lipids, fatty acids, and small molecules such as amino acids, ketones, and glycolysis metabolites [20]. Age, sex, and statin use were included as covariates in all analyses. The data consists of 193,627 (53.8%) females and 166,281(46.2%) males, and the age of participants ranged from ∼40 to ∼73 years old. The distribution of DPW across sex is shown in supplementary Figure F1. Statin use was included as a covariate since many of the predictors are lipid-related, and statins have a profound effect on lipid measures in most people [21]. For this analysis, only first-instance measures were included. Primary ML analysis included only the subset of European-ancestry, unrelated subjects, with subjects of other ancestries reserved for a validation set.

### Outcomes

The primary outcome of interest for prediction from these biomarkers is the number of alcoholic Drinks Per Week (DPW), a quantitative variable. Details of the derivation of this metric in the UKB are described in the supplementary section S1. DPW ranged from 0 to 168 drinks, with a median of 8.6 drinks. Heavy alcohol use, a dichotomous outcome, is defined as consuming ≥15 per week for males or ≥8 per week for females, as defined by the National Institute on Alcohol Abuse and Alcoholism [22]. Additional outcomes investigated included height (Field 50), body mass index (BMI, Field 21001), body fat percentage (BF%, Field 23099), and major depressive disorder (MDD) symptom sum (Category 138). These outcomes were included to serve as benchmark comparisons against which the performance of biomarkers for predicting DPW could be usefully compared. Height was chosen since it is a quantitative measure, stable in adulthood, and any prediction with current biomarker measurements is likely due to biological processes that occurred in the past. BMI and BF% are quantitative anthropomorphic traits that are associated with disease risk and all cause mortality and have a complex relationship with alcohol consumption. In contrast to height, current BMI and BF% represent a mixture of recent and previous biological processes. Finally, major depression symptom count was chosen as an example pseudo-quantitative ordinal variable for a psychiatric disorder.

### Pipeline for ML and GWAS analysis

Figure 1 provides an overview of the data analysis pipeline including data processing, ML methods, GWAS, and post-GWAS analyses. High-performance ML models provide predictions that contribute significantly toward evaluating and interpreting the results as well as subsequent stages of GWAS, heritability estimates, and genetic correlations. In this analysis, we examine seven DPW sets: unfiltered, filtered, and five predicted sets.

**Figure 1.**
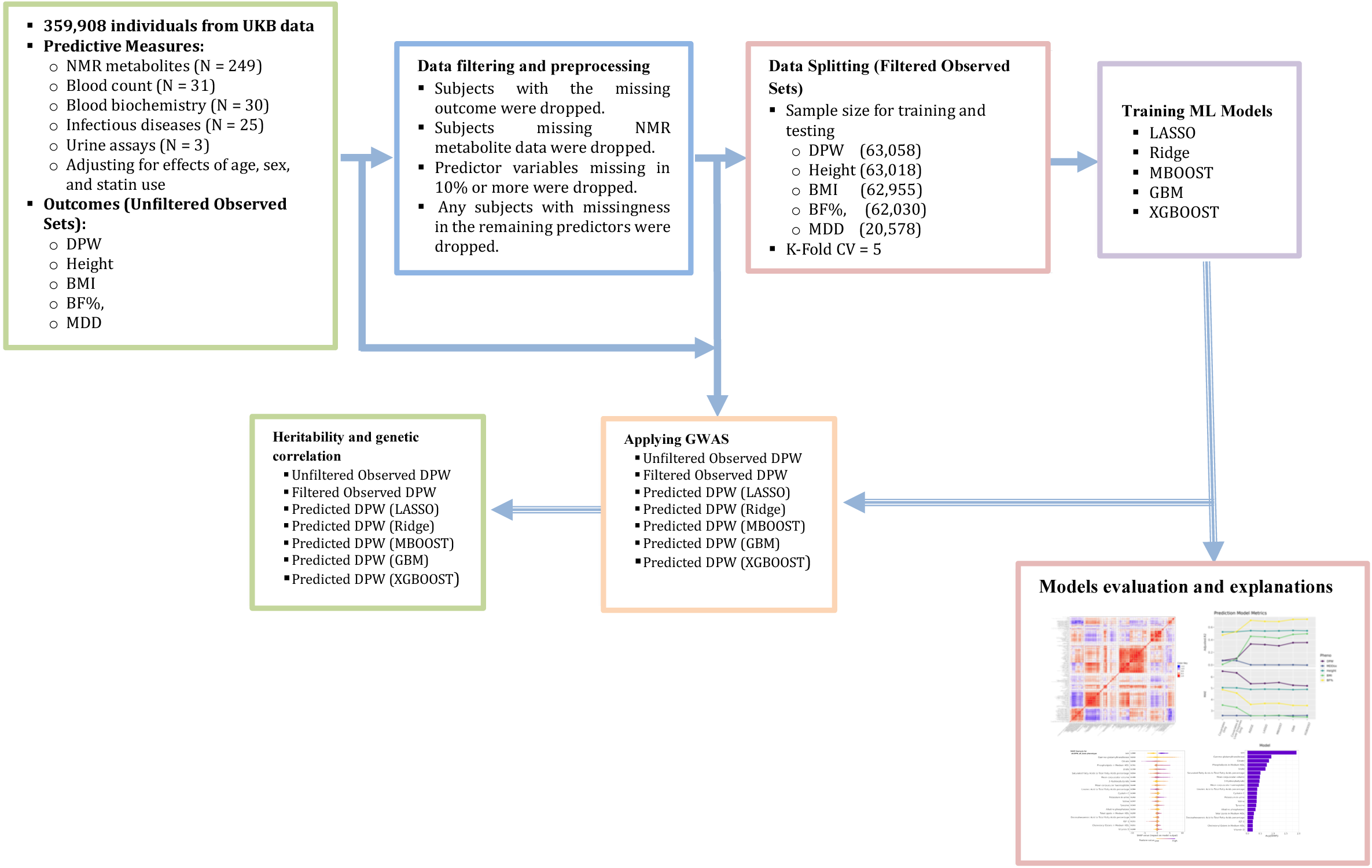
The pipeline is used to train ML models and perform GWAS analysis. The pipeline consists of seven stages. The stages include 1) selecting biomarker variables that are necessary for phenotype prediction, 2) data preprocessing and filtering, 3) data splitting for ML training, 4) training five ML models, 5) ML models evaluation and explanation to provide insight into the role of biomarkers toward final predictions, 6) applying GWAS to the observed and predicted phenotypes, 7) post-GWAS analysis (heritability and genetic correlation) to evaluate the gene discovery capability of ML predictions.

### Data Preparation, demographics, and prevalence

All phenotype predictions and downstream genetic analyses were performed using 359,980 unrelated participants of European ancestry from the UK Biobank. Details of sample filtering for genetic analyses have been previously described [23]. Within the working subset, DPW, Height, BMI, and BF% had little missing data with 359,564 (DPW), 359,118 (height), 358,720 (BMI), and 353,397 (BF%) participants with observations. However, only 117,560 participants had major depressive disorder (MDD) symptom scores since ∼157k subjects completed the mental health questionnaire [UKB Category 138]. In contrast, Nuclear Magnetic Resonance (NMR) based biomarkers [category 220] measures were available in 118,019 participants with 112,767 having complete NMR data.

### Data filtering

Primary prediction and evaluation were restricted to unrelated participants of European ancestry since ancestry may influence some biomarkers. After filtering relatives and participants of non-European ancestry, 82,750 participants remained with complete NMR measures, independent of outcome phenotype. Complete data was retained for 63,058 (DPW), 63,018 (height), 62,955 (BMI), 62,030 (BF%), and 20,578 (MDDsx) subjects. Four independent groups of non-European ancestry participants, as defined by panUKB [24], were used as holdout sets for replication. Four data filtering stages were used in this work. These filtering stages are necessary to prepare data for training. More details about data filtering and sample counts are given in the supplementary Table T3.

### Data Splitting

Data splitting involves dividing the dataset into different subsets intended for training, validation, and testing. The whole dataset is initially separated into a test set and a training set, using a 20% test and 80% training division. Further data splitting within the training set is required to determine the optimal ML model for each implementation. This is done using K-Fold Cross-Validation (CV), a popular technique in machine learning and statistical modeling that evaluates a model’s predictive performance and generalizability. The supplementary Table T3 shows details of the sample counts resulting from the data splitting process.

### ML Evaluation, and Explanation

Five ML models were implemented to predict each outcome, including LASSO [25], ridge regression [26], Gradient Boosting Machines (GBM) [27], Model Boosting (MBOOST) [28], and Extreme Gradient Boosting (XGBOOST) [29]. Models were fit in R (version 4.1.1), using packages glmnet [30], mboost [31], gbm [32], and xgboost [33]. Model optimization parameters are described in the supplementary section S2. To evaluate ML model performance, we used Mean Absolute Error (MAE), Mean Squared Error (MSE), Adjusted R-squared, Accuracy, F1 score, Sensitivity (Recall), Specificity, Positive predictive Value (Precision), and Negative Predictive Value. For the ML model explanation, we used feature importance [34] and SHapley Additive exPlanations (SHAP) [35] [36]. More details about model evaluation and explanations are given in supplementary section S3.

### GWAS in Observed and Predicted Traits

GWAS analyses were performed using BGENIE (version 1.3) [37] on the observed and predicted outcomes from all five predictive models for each phenotype. Standard genotype filtering was performed including removing variants with Minor Allele Frequency (MAF) < 0.5%, INFO score < 0.8, and Hardy-Weinberg Equilibrium (HWE) p-value < 10−6. All GWAS included age, biological sex, and first 20 ancestry principal components as covariates. Primary analyses were restricted to unrelated individuals of European ancestry. Due to small sample sizes (168-1248), no GWAS for h^2^ and r_G_ were performed for the non-European ancestry groups as defined by panUKB.

### Heritability and Genetic Correlation Analyses

Genome-wide Complex Trait Analysis (GCTA, version 1.93.2) [38] was employed for computing heritabilities and genetic correlation (r_G_) between observed and predicted DPW using measured genotypes within the same individuals where DPW was measured. Additionally, LDSC (version 1.0.1) [39] [40] was employed to assess heritabilities and r_G_ between observed and predicted scores using GWAS summary statistics. LDSC allows for the estimation of r_G_ in either independent or overlapping samples by leveraging a reference set of genetic correlations as measured by linkage disequilibrium.

## Results

### Non-independence of biomarkers

Correlation, cluster, and principal component (PC) analyses of the biomarker variables showed extensive non-independence and clustering as illustrated in Figure 2. The first ten PCs explained at least >0.01 of variance each and 0.856 of the variance in aggregate. This pattern is notable since there may be many alternative proxy variables to those that may be retained in the best-fitting ML model. Creating a dendrogram for these biomarkers is beneficial in representing hierarchical relationships among the biomarkers based on their similarities. Biomarkers that are close to each other on the dendrogram are more similar, indicating potential relationships. A dendrogram plot of all biomarkers used to predict DPW is available in the supplemental Figure S2.

**Figure 2.**
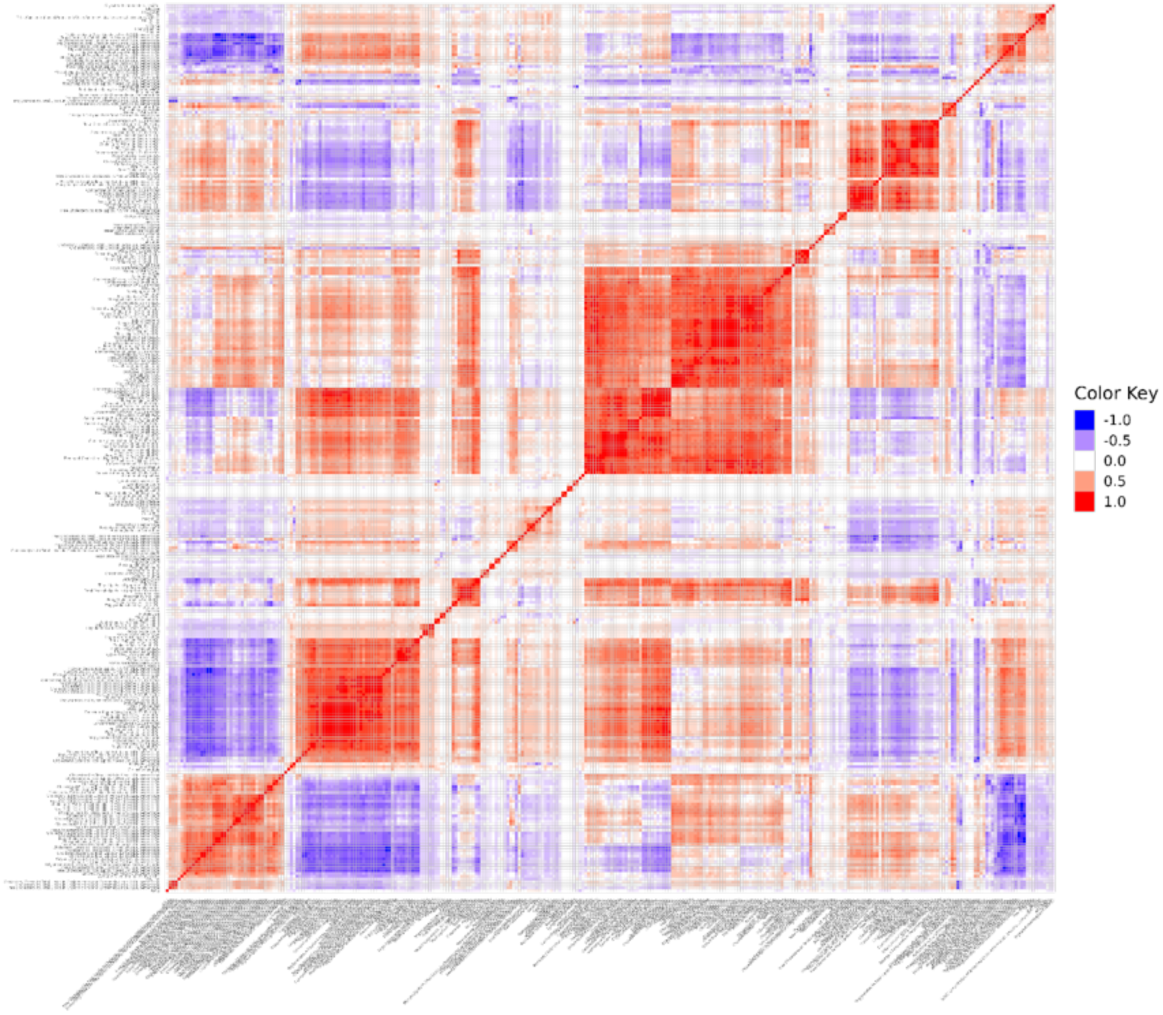
A heatmap of the biomarker variables shows significant correlations, indicating a lack of independence and a tendency to cluster. The observation of extensive correlations suggests there are alternative proxy variables that could potentially be included in the best-fitted ML model.

### ML-based prediction performance

Across five phenotypes, we performed three sets of analyses including (1) a base linear regression model limited to only age, sex, and statin use as predictors, (2) an expanded model adding four liver enzymes (alkaline phosphatase (ALP), ALT, AST, and GGT) to the base model, and finally (3) five ML approaches which included the full set of covariates plus 338 biomarkers. The base model accounted for 0.071 of the variation in DPW. The addition of liver enzymes increased the correlation with observed and predicted DPW to an adjusted R^2^ of 0.105. The five ML approaches utilizing the full set biomarkers increased the adjusted R^2^ to between 0.304 and 0.356. Comparatively, the addition of biomarkers to height prediction did not significantly improve performance in comparison to the base model. In contrast, the inclusion of biomarkers improved BMI and BF% predictions. Finally, MDD symptoms were not well predicted across any of the models tested. A summary of the performance as assessed by MAE and adjusted R^2^ across the 5 phenotypes, 3 models, and 5 ML approaches is shown in Figure 3.

**Figure 3.**
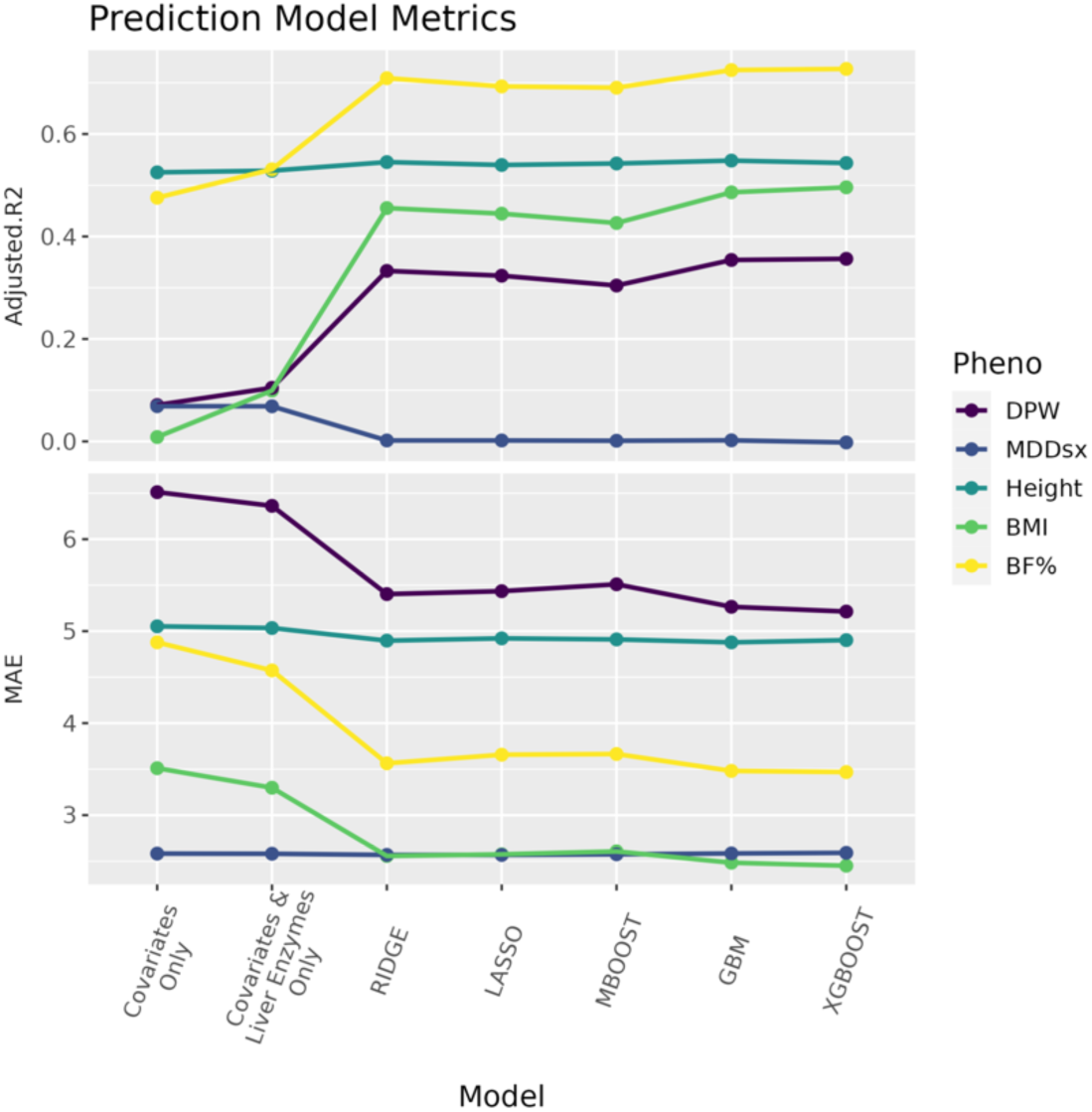
ML model evaluation by Mean Absolute Error (MAE) and adjusted R-squared (R²), for predicting DPW alongside four reference phenotypes. This assessment includes three models: 1) a linear regression model retained only on age, sex, and statin use; 2) a linear ML model trained on four additional liver enzymes (AlkPhos, ALT, AST, and GGT); and 3) five machine learning methods applied to the complete set of covariates alongside 338 biomarker features.

Further details for ML assessment across different phenotypes in terms of adjusted R^2^ and MAE are shown in supplemental Tables T4 and T5, respectively. Inspection of the distributions of observed and predicted DPW showed differences beyond MAE and adjusted R^2^. As shown in Figure 4, the density of DPW predicted by XGboost is the most similar to the observed density, by visual inspection.

**Figure 4.**
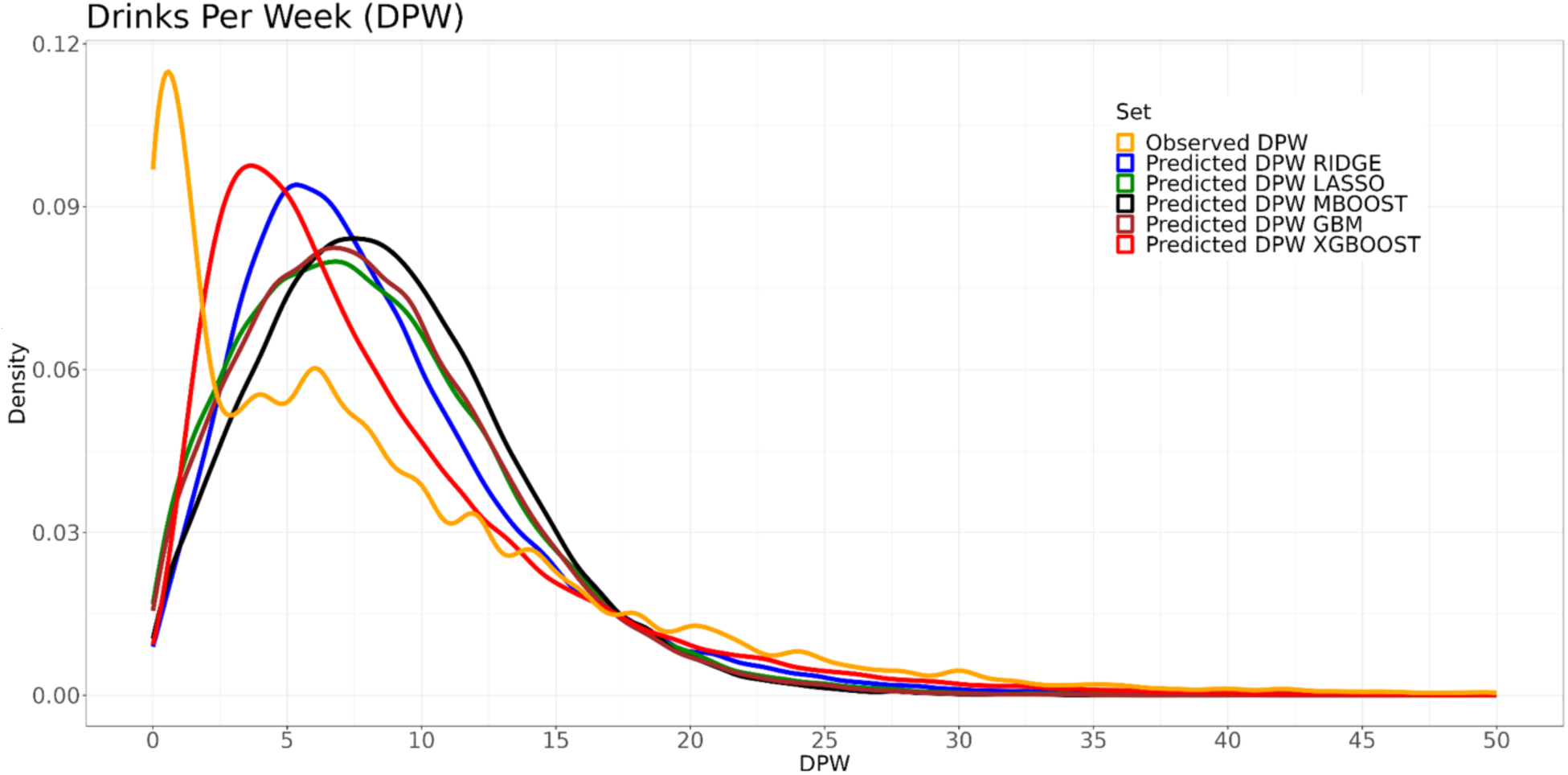
Density plots for observed DPW against predicted DPW across five ML methods: LASSO, Ridge, GBM, MBOOST, and XGBOOST.

### Heritability results

The subset of participants with NMR measures showed higher observed DPW heritability (h^2^=0.0748) than the unfiltered set of participants (h^2^= 0.0643). For the predicted measures, all ML models showed increased h^2^ (delta h^2^ 0.0591-0.0896) compared to the unfiltered set with the exception of XGBOOST (delta h^2^=0.0133) as shown in Figure 5A. More details are given in the supplementary Table T6.

**Figure 5.**
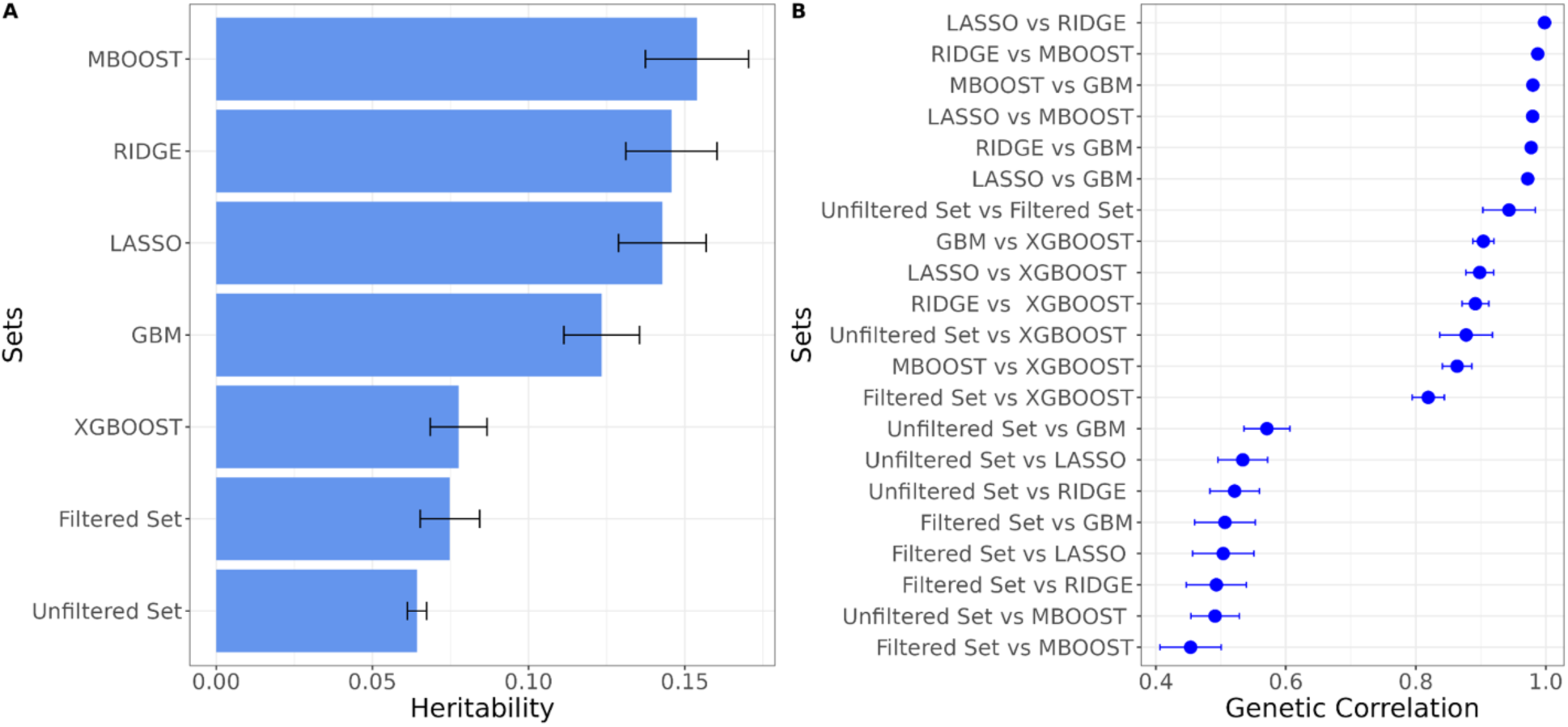
LDSC estimated A) heritabilities and B) genetic correlations.

### Genetic correlations between observed and predicted

In addition to evaluating ML predictions using phenotype correlations, datasets with measured genome-wide genotypes allow the estimation of genetic correlations (r_G_) as shown in Figure 5B. The unfiltered versus filtered sets showed a r_G_ below unity of 0.9434, as expected. Overall, the ML models with increased h^2^ above observed DPW showed lower r_G_. DPW predicted by XGBoost showed both the closest h^2^ to observed as well as the highest r_G_ (0.877) of any of the ML models while MBOOST showed the lowest performance (r_G_=0.4911). The pairwise r_G_ comparisons among the non-XGBoost ML models (LASSO, RIDGE, MBOOST, and GBM) were similar (r_G_ > 0.97). r_G_ between XGBoost and other methods were still high, albeit attenuated (r_G_ 0.863-0.903). While XGBoost predictions were the most similar to observed DPW (similar h^2^ and highest r_G_), the method is more complex to implement and requires additional compute resources (benchmarking not shown). LDSC estimated genetic correlation values for observed and predicted DPW are shown in the supplementary Table T7.

### Assessing predicted heavy use using thresholds applied to predicted values

Predicted DPW was used to classify participants as heavy drinkers based on sex-specific WHO/CDC thresholds, which are 8 and 15 DPW for females and males, respectively. For each of the five ML methods, several classification metrics were calculated, including accuracy, sensitivity, specificity, positive predictive value (PPV), negative predictive value (NPV), and F1 score. The accuracy of classifying all individuals, regardless of sex, was lower for DPW>8 (69.9- 85.6%) in comparison to DPW>15 (86.0-91.9%) across models. For sex specific classifications, all ML methods showed high specificity for both males (89.9%-94.3%) and females (80.5- 93.0%) but moderate sensitivity. As shown in Figure 6, XGBoost performed better as measured by PPV, F1 score, and sensitivity. Sex specific PPVs were relatively low (PPV_male_=63.5%- 81.5%, PPV_female_=49.8%-77.5%) while NPVs were higher (NPV_male_=82.5%-91.2%, NPV_female_=84.6%-90.9%). The full details of performance metrics are given in the supplementary Tables T10 and T11.

**Figure 6.**
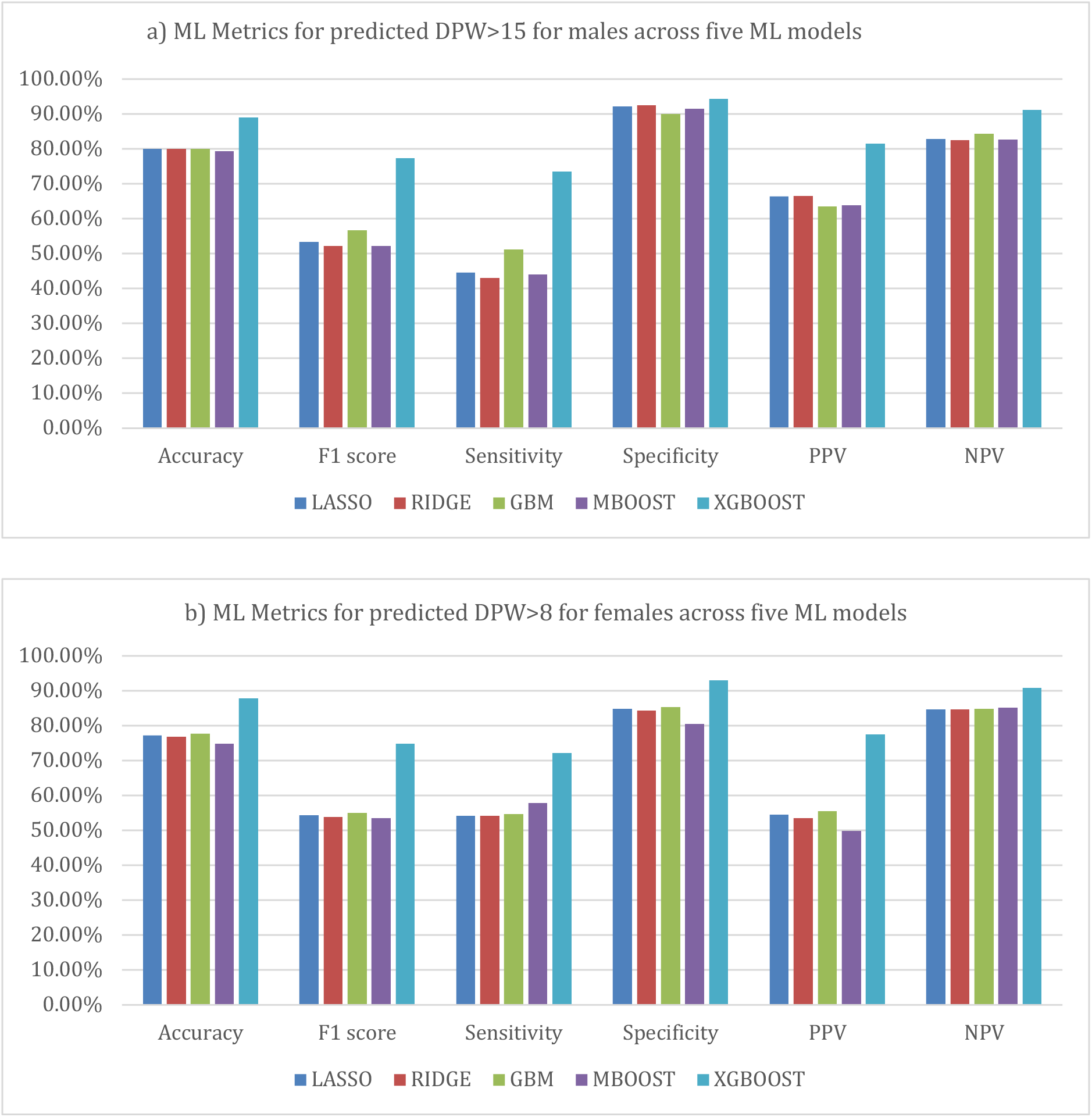
Summary of gender specific heavy drinker classification performance using predicted DPW by six evaluation metrics including accuracy, F1 score, sensitivity, and specificity, PPV, and NPV. For males (Figure 6a), DPW greater than 15 is considered heavy drinking while for females (Figure 6b), DPW greater than 8 is considered heavy drinking. These metrics summarize the prediction performance of five ML methods for classifying heavy drinkers in males and females.

### Replication in participants of non-European ancestry

We also assessed the prediction performance in five independent holdout sets representing participants with genetic ancestry similar to reference population sets including African (AFR, N = 706), Admixed Americas (AMR, N = 168), central Asia (CSA, N = 1248), east Asia (EAS, N = 420), and the Middle East (MID, N = 242). Sample sizes for each replication set are based on participants with complete NMR and DPW data. Replication was limited to the XGBoost model due to its higher performance across multiple prediction metrics. MAE across these ancestry groups was similar to the EUR test set (MAE = 5.2, range = 4.3 - 5.1). In comparison to the EUR test set, accuracies, specificities, and NPV were higher for the non-EUR sets, while sensitivity and PPV were generally lower.

### ML model interpretation

While a variety of ML approaches are available to predict outcomes like DPW, interpreting the contribution of different biomarkers is important for several reasons including evaluating plausibility, generalizability, and identifying confounders or artifacts. Figure 7 shows the top 20 biomarkers by feature importance that contribute to the prediction of DPW across the boosting-based ML models. The markers are arranged in descending order and the scores of each marker represent the median across ML models. In addition to examining feature importance, SHAP analyses help in interpreting the local and global contributions of each variable to the prediction of a given ML model. Details regarding the estimation and interpretation of SHAP values are given in supplemental section S3.3. Figure 8 shows the Shapley values from the best-performing XGBOOST model for DPW and includes the top 20 biomarkers. The cloud of points shown for each predictor represent each observation (in this case participants) and the relative importance of that biomarker for predicting the outcome for that observation. Higher absolute SHAP values indicate a stronger effect of the biomarker on the prediction, while values towards zero indicate a less of an impact. For instance, males tend to have higher DPW compared to females, and sex shows significant variance, leading to the highest average absolute SHAP values. Another example is citrate: as citrate levels increase (bluer color), SHAP values become negative (indicating low DPW), while as citrate decreases (yellower color), SHAP values turn positive (indicating high DPW).

**Figure 7.**
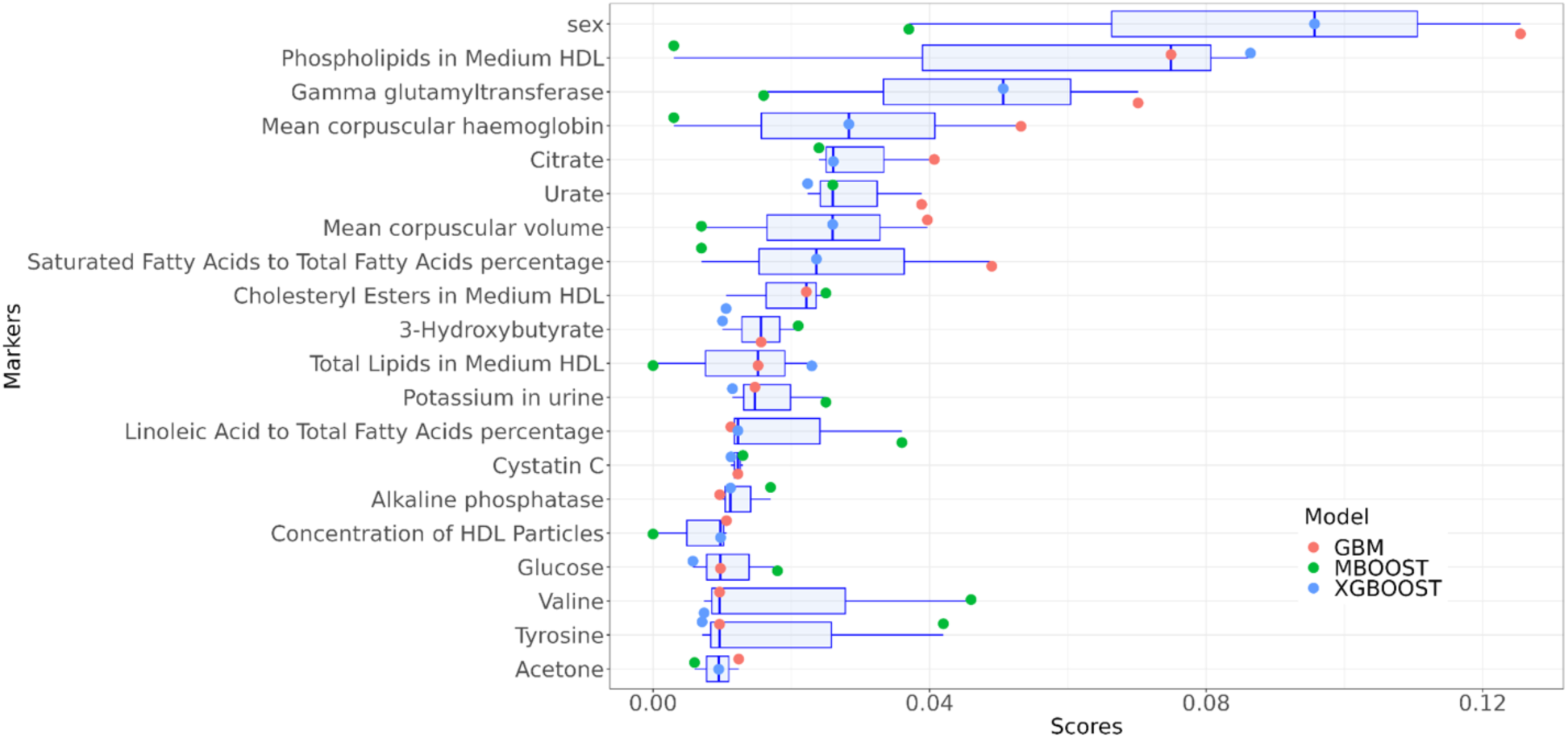
Top 20 biomarkers influencing DPW prediction across boosting-based machine learning models. These markers are listed in descending order, with their respective scores representing the median feature importance across all boosting ML models.

**Figure 8.**
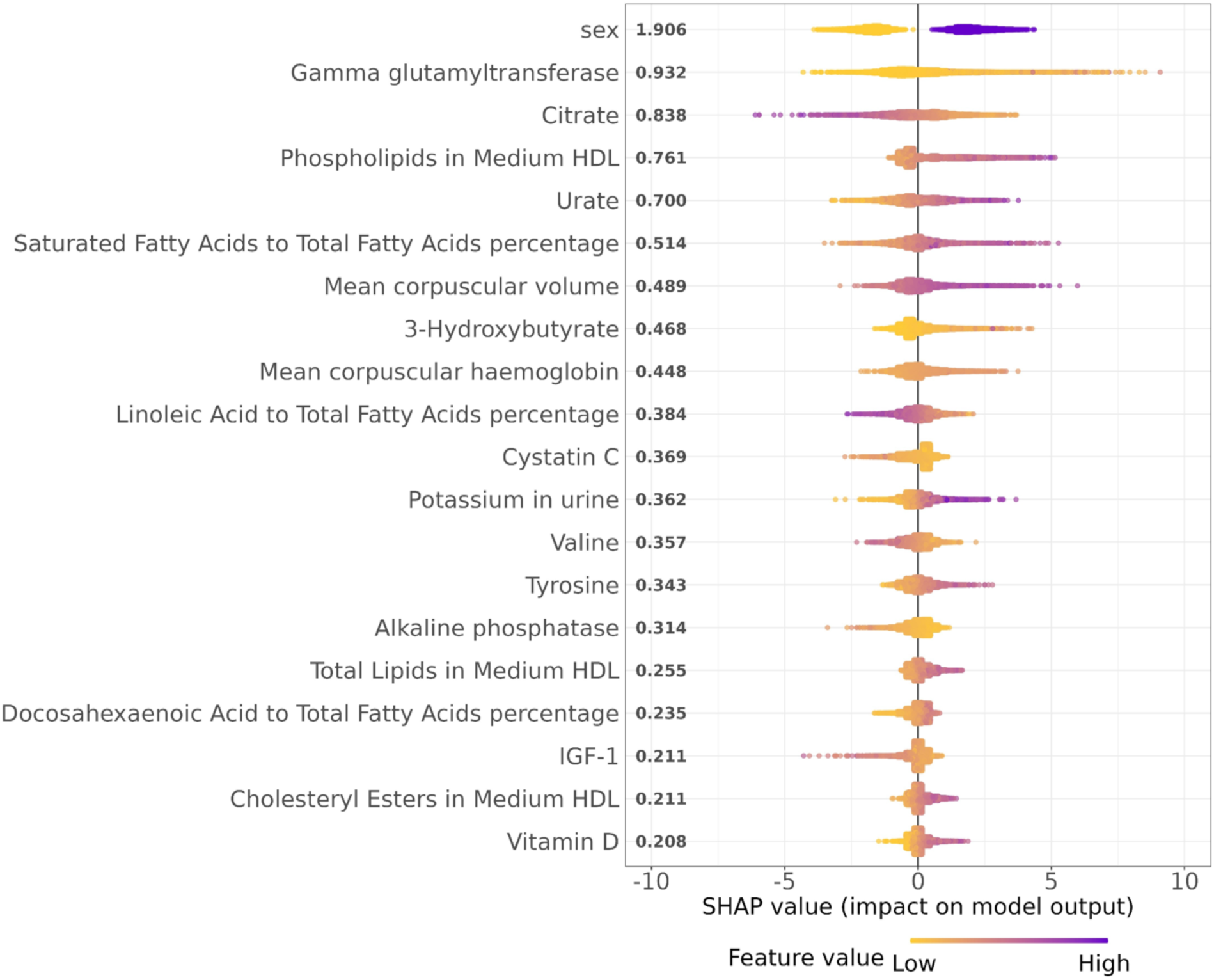
Shapley values for the top 20 biomarkers from XGBOOST based prediction of DPW. Each data point corresponds to a measurement from an individual participant. Blue dots signify higher values, while yellow dots represent lower values. A higher SHAP value indicates a positive impact on the DPW, whereas a lower SHAP value suggests a negative impact.

## Discussion

To our knowledge, this study represents the first of its kind to utilize a wide array of biological markers for ML prediction of alcohol consumption in an agnostic fashion. In total, five different ML models were applied to predict DPW and demonstrated strong predictive performance, as measured by multiple standard metrics (R_adj_^2^, MAE, MSE) as well genetic based measures including heritability (h^2^) and genetic correlation (r_G_), both in hold-out test sets of European ancestry and in independent test sets of various diverse ancestries. This proof-of-concept experiment confirms the viability of ML for predicting alcohol consumption accurately from lab-based metrics, suggesting potential for future clinical screening applications.

### Implications of DPW Prediction

#### Important predictors

This analysis identified various biological measures associated with DPW and useful for predicting this outcome, including some which have been previously identified in the literature, such as sex, HDL, and GGT. The relationship between sex and alcohol consumption is complex and influenced by various factors including biological differences, social, cultural, and psychosocial factors, as well as hormonal influences [41]. Alcohol consumption has been associated with changes in lipid metabolism, including effects on HDL cholesterol levels. Moderate alcohol intake has also been linked to an increase in HDL cholesterol. However, the relationship between alcohol and HDL is complex, and the effects may vary depending on factors such as the type and amount of alcohol consumed, individual characteristics, and overall health [42]. Alcohol is known to increase GGT levels, which is considered a sensitive marker for alcohol-induced liver damage. Elevated levels of GGT in the blood can be associated with various conditions, including liver diseases, biliary obstruction, and certain medications. One of the significant factors contributing to elevated GGT levels is excessive alcohol consumption [43]. As shown in this experiment, age, sex, and liver enzymes explain a modest amount of variation in DPW (R_adj_^2^∼0.11).

#### DPW predictions for AUD screening

Improved screening to identify heavy alcohol drinkers is important since progression to AUD and related health and psychosocial problems is a process that occurs over time. Identification of heavy drinking using objective biomarkers may represent an additional complementary tool to established screening instruments. Interventions and treatments are likely more effective in at-risk individuals prior to progression to AUD or when AUD is still mild [44]. Additionally, most people with AUD do not seek treatment. Identifying and offering empirically supported treatment to non-treatment seeking individuals with AUD offers the potential to reduce the significant negative personal, societal, and economic impact of AUD [45].

While we demonstrated the current predictions are useful to screen for heavy drinking, there is a significant opportunity for improved performance for low to moderate consumption. Dramatic improvements using other ML methods and the same set of biomarkers are unlikely and will require additional predictors. Improved prediction at the lower end of the scale could allow healthcare professionals to track changes in alcohol consumption over time and allow earlier interventions. While individual level predictions may have errors, biomarker-based predictions are useful in genetic research as effective sample size is increased and may have applications in population-level research on alcohol consumption patterns. As predictions improve, this information will be valuable for public health initiatives, policy development, and the allocation of resources for prevention and treatment programs.

### Machine Learning Considerations and Limitations

#### Distribution of alcohol consumption and implications on ML predictions

One potential challenge to ML models is non-normal distributions as seen in DPW, where most of the participants are relatively light drinkers (majority class), but there are a significant number of heavy drinkers (minority class), resulting in a long tail. This can lead to biased models that perform well on the majority class but poorly on the minority class. Right-skewed distribution (RSD) causes high specificity values since the majority of the data is concentrated toward light drinkers. At the same time, RSD causes lower sensitivity because the tail of the distribution (heavy drinkers) contains fewer data points. Boosting techniques such as MBOOST, GBM, and XGBoost can be adjusted to handle class imbalance by focusing more on the minority class and this is an important area for future development.

#### Evaluating predictions

Although five different ML models were evaluated, the performances were similar across methods and outcomes as assessed by R_adj_^2^, MAE, and MSE. However, genetics-informed evaluation of model performance showed XGBoost to perform the best with the most similar h^2^ and highest r_G_ with observed DPW. These analyses also demonstrated that while the subset of participants with NMR measures showed higher heritability (h^2^=0.0748) when compared to everyone with measured DPW (h^2^=0.0643), the difference was not significant and the genetic correlation was high (r_G_=0.94). The high observed r_G_ supports the use of predictions even if the variance explained is modest (R_adj_^2^∼0.35).

#### ML predictions using highly correlated data

We also demonstrate that all ML models tested are able to produce predictions from a large set of highly non-independent predictors (Figure 2). Due to this non-independence, caution in interpretation of variable importance across models is warranted. While the LASSO model performs variable selection, all predictors are retained in the final models for the other methods and their relative importance can vary greatly. Given the non-independence of predictors, the relative rank of predictors across models is not necessarily meaningful. However, many of the most important predictors by SHAP analysis have a known relationship to alcohol consumption including the top three; sex, phospholipids in medium HDL, and GGT. Additional details of the relationship between correlated predictors are described in the supplementary Figure S2. A full list of all SHAP values for all biomarkers including covariates is shown in the supplementary Table T12 ranked in descending order.

Figure 8 illustrates that different markers display varying SHAP value distributions, revealing unique insights into the role of markers in predicting alcohol consumption. For example, markers that are highly connected with high alcohol consumption are GGT enzyme (high values), followed by mean corpuscular volume (high values), saturated fatty acid to total fatty acid percentage (high values), phospholipid in medium HDL (high values), etc. In contrast, markers that are highly connected with low alcohol consumption are citrate (high values), followed by GGT enzyme (low values), IGF-1 (high values), etc. While some markers have bidirectional effects on alcohol consumption (low vs high), others have unidirectional effects. For example, low values of phospholipid in medium HDL are not important in predictions compared to high values which show a potential relationship with high alcohol consumption.

#### MAE and prediction interpretation

While metrics such as R² are good summaries of overall model performance, they are open to subjective interpretation. Importantly, how the predictions will be used can influence interpretation. The observed MAE of 5.2 in this study implies prediction at the lower end of the DPW scale (0-5) is less useful. However, this MAE is less problematic at the higher end of the scale (DPW>20). We demonstrated empirically that predicted DPW with this level of imprecision is still useful in classifying heavy drinkers using established thresholds. The observed sensitivities and specificities may be useful in some screening strategies.

### Limitations of UKB

The UK Biobank represents an important resource for biomedical research, but research findings using this sample do come with several important limitations for generalizability. Participants in the UKB are, on average, healthier and of higher socio-economic status than the general UK population. Of particular relevance for genetic analyses, participants are largely of white, British ancestry. Furthermore, all participants are UK residents and cultural, environmental, legal, and healthcare system differences must be considered when attempting to generalize results to populations across different geographical regions.

## Conclusion

The development of efficient tools for measuring and/or predicting AC is important for both research and future clinical applications. The combination of ML and biomarkers presents an opportunity to predict alcohol consumption patterns. These predictions could be particularly valuable in large-scale genetic studies lacking self-report or structured interviews related to AC but with available biomarkers. This study demonstrates that ML methods can predict AC using a large set of biomarkers and that genetic analyes are useful in evaluating prediction performance beyond standard approaches using subsets of the data as holdouts. The findings indicate that all ML models produced useful predictions of AC in the context of classifying heavy drinking.

Genetic analysis revealed similar heritability estimates and high genetic correlation between observed and predicted DPW using XGBOOST in this sample. By design, the ML methods used here yielded interpretable models where the relationship between important biomarkers and AC can be further evaluated. Finally, the ML models produced by this work for predicting DPW and/or heavy drinking may provide the basis of complementary tools for alcohol related screenings in clinical applications with continued development.

## Data Availability

All data produced in the present work are contained in the manuscript

## Acknowledgments and Disclosures

This research has been conducted using the UK Biobank Resource application number 30782. MH, REP, BTW, and AEG were supported by NIH P50AA022537, REP, BTW, and AEG by NIH R01MH125938, REP by The Brain & Behavior Research Foundation NARSAD grant 28632 P&S Fund, AEG by NIH 1K01AA031748-01 and ECP by 1R01DA054313-01A1.

The authors report no financial interests or potential conflicts of interest.

