## Supplementary Material for "Machine Learning Approaches to Predict Alcohol Consumption from Biomarkers in the UK Biobank"

Mohammed F. Hassan[*1,2], Amanda Elswick Gentry[*1,2], Elizabeth C. Prom-Wormley [1,3], Roseann E. Peterson[4,1], Bradley T. Webb[5,1]
[*]Joint-First Authors
[1]Virginia Institute for Psychiatric and Behavioral Genetics, Virginia Commonwealth University, Richmond, Virginia, USA;
[2]Department of Psychiatry, School of Medicine, Virginia Commonwealth University, Richmond, Virginia, USA;
[3]Department of Epidemiology, School of Public Health, Virginia Commonwealth University, Richmond, Virginia, USA;
[4]Department of Psychiatry and Behavioral Sciences, Institute for Genomics in Health, SUNY Downstate Health Sciences University, Brooklyn, New York, USA;
[5]GenOmics and Translational Research Center, RTI International, Research Triangle Park, North Carolina, USA

### 1. Supplementary Section S1: Definition of Drink Per Week in UK Biobank

Drinks Per Week (DPW) calculation in UK Biobank (UKB) data is quantified as the average number of alcoholic drinks consumed by a participant per week. UKB collects this data through self-reported questionnaires where participants are asked about their typical drinking habits, including the frequency and quantity of alcohol consumption. The details of the DPW calculation can be summarized as follows:

- **Questionnaire Data**: Participants are asked how often they drink alcohol and how many drinks they typically consume on each occasion. The questions usually cover different types of alcoholic beverages, such as beer, wine, spirits, etc.
- **Frequency of Drinking**: This data includes:
    - Daily
    - 3-4 times a week
    - Once or twice a week
    - 1-3 times a month
    - Rarely
    - Never
- **Quantity Per Occasion**: Participants indicate how many drinks they typically consume per drinking session, which can vary by drink type such as number of glasses of wine.

The formula used for the calculation of DPW is defined as

$$DPW = (\,Frequency\ of\ \ drinking\,) \times (\,Number\ of\ \ Drinks\ per\ Occasion\,) \qquad (1)$$

### 2. Supplementary Section S2: Review of machine learning methods

#### 2.1. LASSO and Ridge Regressions

LASSO stands for Absolute Shrinkage and Selection Operator. It is an L1 regularization technique to estimate the relationship between independent variables and the dependent variable. For N samples, the response (dependent) variable ($y_i$) is approximated by a linear combination of independent variables [1]

$$\bar{y}(x_i) = \beta_0 + \sum_{j=1}^{p} x_{ij}\beta_j. \tag{2}$$

The model is parameterized by the vector of regression weights $\beta_0, \beta_1, \ldots, \beta_p$ that are optimized during the fitting process and $\bar{y}_i$ is the estimated dependent variable. To estimate the fitted line, the variable coefficient $\beta_0, \beta_1, \ldots, \beta_p$should be optimized according to the following loss function

$$arg_{\beta_0\beta_j}\, min\left\{\frac{1}{N}\sum_{i=1}^{N}\left(y_i - \beta_0 - \sum_{j=1}^{p} x_{ij}\beta_j\right)^2\right\} \tag{3}$$

In LASSO, the loss function is modified by adding a regularized term to the loss function which is the sum of the absolute value of the coefficients as defined in the following objective function

$$arg_{\beta_0\beta_j}\, min\left\{\frac{1}{N}\sum_{i=1}^{N}\left(y_i - \beta_0 - \sum_{j=1}^{p} x_{ij}\beta_j\right)^2\right\} + \lambda\sum_{j=1}^{p}\left|\beta_j\right| \tag{4}$$

To get the best-fitted line, the cost function should be optimized by forcing the coefficient variables to zero and thus directly performing feature selection where λ is a tuning parameter that controls the strength of the penalty.

Ridge regression [2] is considered as L2 regularization, the added regularized term to the loss function is the sum squares of the coefficients as defined in the following objective function

$$arg_{\beta_0\beta_j}\, min\left\{\frac{1}{N}\sum_{i=1}^{N}\left(y_i - \beta_0 - \sum_{j=1}^{p} x_{ij}\beta_j\right)^2\right\} + \lambda\sum_{j=1}^{p}\beta_j^{\,2} \tag{5}$$

Although LASSO and Ridge regressions are built using similar structures, they have many key differences. LASSO can perform feature selection by shrinking some coefficients to zero, while Ridge regression does not. In terms of model complexity LASSO can create simpler, more interpretable models by selecting a subset of the features. Ridge regression typically includes all features but with smaller, more balanced coefficients. Regarding handling multicollinearity both methods handle multicollinearity, but Ridge is more effective when all predictors are relevant as it keeps all features. In terms of performance Ridge regression may perform better when dealing with highly correlated features, while LASSO may be more useful when dealing with high-dimensional data with many irrelevant features.

Key Parameters for LASSO and Ridge [3]

- Lambda (λ): This is the regularization parameter. Higher values of λ lead to more regularization, which means that the coefficients can be reduced to zero, effectively performing feature selection and often determined using cross-validation.
    - minimum λ for LASSO regression = 0.000256
    - minimum λ for Ridge regression = 0.2558.
- Alpha: Determines the type of regularization
    - alpha = 1: Lasso regression.
    - alpha = 0: Ridge regression.
    - 0 < alpha < 1: Elastic Net (combination of Lasso and Ridge).
- Family: The type of model being fit
    - "Gaussian": Linear regression.
    - "binomial": Logistic regression.
    - "poisson", "multinomial.
        - Family = "Gaussian".
- Intercept: A boolean value indicating whether to calculate the intercept for the model. If set to False, no intercept will be used in the calculations.
    - Intercept = True.
- Standardize: A boolean value, when set to True, standardizes the regressors before fitting the model. Standardization means that the mean will be 0, and the standard deviation will be 1.
    - Standardize = True.
- Max Iter: The maximum number of iterations for the optimization algorithm. If the algorithm does not converge within this number of iterations, it will stop.
    - Max Iter= 100.
- Tolerance: The tolerance for the optimization. It determines when the optimization should stop if the improvement in the cost function is below this threshold.
    - Tolerance = 1e-7.

### 2.2. MBOOST, GBM, and XGBoost

MBOOST stands for model boosting [4], GBM for gradient boosting [5], and XGBOOST [6] for extreme boosting. These algorithms refer to different implementations of boosting algorithms used in machine learning for regression and classification tasks. In general, they build on similar structures by combining the outputs of several weak models to create a strong predictive model. The primary idea is to sequentially apply the weak models to adjust the weights of incorrectly classified instances and focus more on the difficult cases in subsequent iterations.

MBOOST is an R package designed for model-based boosting, offering flexible and interpretable boosting methods. MBOOST package supports a wide range of base learners and loss functions and is intended to provide interpretable models, making it easier to understand the impact of individual predictors. However, it may be slower and less efficient compared to other implementations, particularly for large datasets.

Key Parameters for MBOOST [4]

- Base Learners: Define the type of model used in each boosting iteration. Some common base learners and their defaults include:
    - bbs (B-splines)
    - bols (Linear effects)
    - btree (Decision stumps/trees)
        - base learners = "bols", number of base learners = 303.
- Family (Loss Function): The loss function is specified using the family parameter. Common choices for regression include:
    - Gaussian ()
    - Laplace ()
    - Huber ()
        - Family = Gaussian.
- Number of Boosting Iterations (mstop): This can be optimized using cross-validation.
    - mstop = 1000.
- Learning Rate (nu): determines the step size at each boosting iteration. Smaller values lead to slower learning but can result in better generalization.
    - nu = 0.1.

GBM is another R package that provides an implementation of gradient boosting. It is one of the earlier implementations and is widely used for a variety of predictive modeling tasks. It is suited for a range of tasks and includes a variety of options for tuning and regularization. GBM can handle various types of data and tasks. It is robust to overfitting with proper tuning and capable of capturing complex patterns.

Key Parameters for GBM [7]

- Distribution: Specifies the loss function used in the model. For regression tasks, common choices are:
    - "laplace": For regression with a Laplace loss function.
    - "gaussian" is typically used for continuous target variables in regression problems.
        - Distribution = "gaussian"
- n.trees: The number of trees to fit (i.e., the number of boosting iterations). Typically chosen through cross-validation.

- n.trees = 10000.
- interaction.depth: The maximum depth of each tree. It controls how complex the individual trees can be. Larger values lead to more complex trees.
  - interaction.depth = 2
- shrinkage (or learning.rate): The learning rate or shrinkage factor. It controls the contribution of each tree. A smaller value makes the learning process more conservative and reduces the risk of overfitting.
  - Shrinkage = 0.01.
- n.minobsinnode: The minimum number of observations required in a terminal node (leaf). This controls the size of the trees. Larger values can prevent overfitting.
  - n.minobsinnode = 15.
- verbose: Whether to print progress messages during the fitting process.
  - verbose = FALSE.
- bag.fraction: The fraction of the training data used for each iteration of boosting. This can help prevent overfitting by creating a more diverse set of trees.
  - bag.fraction = 0.5.

XGBoost is an R package that provides a highly efficient and scalable implementation of the gradient boosting framework. It is designed for speed and performance, making it a popular choice for machine learning tasks, particularly those involving structured/tabular data. XGBoost is designed to be efficient in both computation and memory usage. It supports parallel processing, making it faster than many other implementations of gradient boosting. It includes L1 and L2 regularizations to prevent overfitting, which can improve the model's generalization capabilities. It can handle missing values within the dataset, using a sparse matrix format and a special handling strategy.

Key Parameters for XGBoost [8]

- General Parameters: These parameters control the overall behavior of the XGBoost model.
  - booster: Type of booster to use.
    - Options: "gbtree", "gblinear", "dart"
      - Booster = "gbtree"
  - nthread: Number of threads to use for training.
    - nthread = 1 (single-threaded)
  - verbosity: Controls the level of messages printed.
    - Options: 0 (silent), 1 (warnings), 2 (info), 3 (debug)
- Booster Parameters: Tree Booster (gbtree and dart)
  - eta (or learning_rate): Step size shrinkage to prevent overfitting.
    - eta = 0.01
  - max_depth: Maximum depth of a tree.
    - max_depth = 15
  - min_child_weight: Minimum sum of instance weight (hessian) needed in a child.
    - min_child_weight = 1

- gamma: Minimum loss reduction required to make a further partition.
  - gamma = 100
- subsample: Fraction of samples used to grow trees.
  - Subsample = 1
- colsample_bytree: Fraction of features used per tree.
  - colsample_bytree = 1
- colsample_bylevel: Fraction of features used per level.
  - colsample_bylevel = 1.0
- colsample_bynode: Fraction of features used per split.
  - colsample_bynode = 1.0
- lambda (or reg_lambda): L2 regularization term on weights.
  - Lambda = 100
- alpha (or reg_alpha): L1 regularization term on weights.
  - alpha = 100
- tree_method: Tree construction algorithm.
  - Options: "auto", "exact", "approx", "hist", "gpu_hist"

- Learning Task Parameters: These parameters are specific to regression tasks.
  - objective: Specifies the learning task.
    - Common options for regression:
      - "reg:squarederror" (default): Mean squared error regression.
      - "reg:logistic": Logistic regression.
      - "reg:pseudohubererror": Pseudo Huber regression.
    - Other advanced options include quantile and Poisson regression.
  - eval_metric: Evaluation metrics for validation data.
    - Options:
      - "rmse": Root Mean Squared Error (default for regression).
      - "mae": Mean Absolute Error.
      - "mape": Mean Absolute Percentage Error.
      - "poisson-nloglik": Negative log-likelihood for Poisson regression.
      - "gamma-nloglik": Negative log-likelihood for gamma regression.
      - "logloss": Log loss.
  - base_score: Initial prediction score (global bias).
    - base_score = 7.524918

### 3. Supplementary Section S3: ML Model Evaluation and Explanation.

#### 3.1. Model Evaluation

In order to compare the performance of machine learning models, we need to select an appropriate tool for a given task (regression or classification), which is considered essential to assess the model's performance and make informed decisions.

For regression tasks [9], there are several evaluation metrics and techniques commonly used to assess the quality of regression models. In this work, we used two methods; the first is the Mean Absolute Error (MAE), which measures the average absolute difference between predicted and actual values. It is less sensitive to outliers compared to some other metrics. MAE is defined as

$$MAE = \left(\frac{1}{n}\right) \times \sum \left| Y_i - \overline{Y}_i \right| \quad (6)$$

where $n$ is the number of data points. $Y_i$ and $\overline{Y}_i$ are actual, and predicted phenotypes, respectively. The second method called Adjusted R-squared is a statistical metric used in regression analysis to assess the goodness of fit of a regression model. It is an improvement over the regular R-squared and takes into account the number of predictor variables in the model. R-squared measures the proportion of the variance in the dependent variable that is explained by the independent variables in a regression model. However, as more independent variables are added to the model, R-squared tends to increase, even if those additional variables do not significantly improve the model's predictive power. Adjusted R-squared addresses this issue by penalizing the inclusion of unnecessary variables in the model. It is calculated using the formula:

$$Adjusted\ R\_Squared = 1 - \left[\left(1 - R\_squared\right) \times \frac{(n-1)}{(n-p-1)}\right] \quad (7)$$

where R-squared is the regular coefficient of determination. n is the number of observations (data points), and p is the number of predictor variables in the model.

For classification tasks [10], different ML metrics are used for evaluation. All these metrics are derived from the confusion matrix. A confusion matrix is a table used to evaluate the performance of a classification algorithm. It summarizes the predicted results of a model by comparing the actual (true) labels with the predicted labels for a set of data. It is especially useful in binary and multiclass classification problems.

The confusion matrix is defined as follows:

| | **Predicted Positive** | **Predicted Negative** |
|---|---|---|
| **Actual Positive** | True Positive ($TP$) | False Negative ($FN$) |
| **Actual Negative** | False Positive ($FP$) | True Negative ($TN$) |

- True Positive ($TP$): The number of instances correctly predicted as positive.
- True Negative ($TN$): The number of instances correctly predicted as negative.
- False Positive ($FP$): The number of instances incorrectly predicted as positive (also called a "Type I error").
- False Negative ($FN$): The number of instances incorrectly predicted as negative (also called a "Type II error").

The following are definitions of the metrics used throughout this work

- Accuracy: Measures overall correctness.

$$Accuracy = \frac{(TP + TN)}{(TP + TN + FP + FN)}.$$

- Positive Predictive Value (Precision): Measures the proportion of correct positive predictions.

$$Postive\ Predictive\ Value = \frac{TP}{(TP + FP)}.$$

- Sensitivity (Recall or True Positive Rate): Measures the proportion of actual positives that are correctly identified

$$Sensitivity = \frac{TP}{(TP + FN)}.$$

- Specificity (True Negative Rate): Measures the proportion of actual negatives that are correctly identified.

$$Specificity = \frac{TN}{(TN + FP)}$$

- $F1\ score$: A harmonic mean of precision and recall, useful when there is an imbalance between positive and negative samples.

$$F1\ score = \frac{2(Precision \times Recall)}{(Precision + Recall)}.$$

- Negative Predictive Value (NPV): Measures the probability that individuals who test negative for a condition truly do not have the condition.

$$Negative\ Predictive\ Value = \frac{TN}{(TN + FN)}.$$

#### 3.2. Model Hyperparameter Tuning

All ML algorithms mentioned above need tuning [11], often referred to as hyperparameter tuning, which is the process of optimizing the hyperparameters during training of a machine learning model to achieve better performance on a specific task or dataset. Properly tuning hyperparameters can significantly improve the generalization performance of a machine-learning model. The tuned parameters for each ML model depend on its internal structure and are learned using different methods. In this work, we used the grid search to find the best hyperparameter values. A grid search is used to exhaustively search through all possible combinations of hyperparameters within the defined search space. It's simple but can be computationally expensive. In this work, the hyperparameters used in the training five ML methods are defined in the Supplementary Section S2.

#### 3.3. Model Interpretability

Machine learning interpretability is crucial when applied to the field of biology to extract meaningful insight [12]. By understanding the overarching process by which the input variables generate why a model makes a certain prediction, researchers can identify relevant phenotypes, molecules, genes, proteins, or pathways that are involved in a particular biological phenomenon. In this work, we used feature importance, a concept commonly used in machine learning and data analysis to understand the relative significance of input variables (blood biomarkers) in predicting a given phenotype. It helps in identifying which markers or covariate variables have the most influence on the model's predictions. Various techniques can determine feature importance depending on a given machine learning algorithm. In models that use L1 regularization (e.g., LASSO), features that have non-zero coefficients after regularization are considered important, while features with zero coefficients are considered unimportant. Boosting algorithms (e.g., MBOOST, GBM, XGBOOST) use decision trees as base models. In decision trees, features that split the data into classes effectively are considered more important. Information gain or Gini impurity is commonly used to evaluate the quality of a split. Features that lead to nodes with a substantial reduction in impurity are considered more important.

SHapley Additive exPlanations (SHAP) [13] is another method that is used both in local and global interpretably of ML. SHAP is a framework for interpreting the output of machine learning models. It is based on cooperative game theory and the concept of Shapley values, which were introduced by Lloyd Shapley. The Shapley value provides a way to fairly distribute rewards among a group of contributors. In the context of machine learning interpretability, SHAP values aim to attribute the contribution of each feature to the prediction made by a model. These values provide insights into the importance of different features in determining the output of a model for a particular instance. The mathematical formula for SHAP analysis can be summarized as follows

$$g(x) = \mu_0 + \sum_{i=1}^{N} \mu_i(g, x), \tag{8}$$

where

- $g(x)$ is the model's prediction for an instance x.
- $\mu_0$ is the base value or the average model output over the training dataset.
- $\mu_i(f, x)$ is the SHAP value for the feature $i$.
- $N$ is the number of features.

The SHAP value for the feature $i$ is calculated as follows

$$\mu_i(g, x) = \sum_{S \sqsubseteq M \setminus \{i\}} \frac{|S|! \times (|M| - |S| - 1)!}{|M|!} \times \left[ g\left( x_s \cup \{i\} - g\left( x_s \right) \right) \right], \tag{9}$$

where

- $M$ is the set of all features.
- $S$ is a subset of features that does not include feature $i$.
- $|S|$ is the number of elements in the subset $S$.
- $g\left(x_s\right)$ is the model prediction for the subset $S$ of features.

- $g\left(x_s \cup \{i\}\right)$ is the model prediction for the subset $S$ of features plus feature $i$.

### 4. Supplementary Table T1: UKB Variables

The UKB is a large-scale biomedical research resource with data from ~500,000 volunteer participants aged between 40-69 years recruited in 2006-2010 from across the UK and contains a rich array of measures including lifestyle and health surveys, EHR derived diagnoses, blood chemistry, genetics, metabolomics, proteomics, imaging, and accelerometer data. With their consent, participants provided detailed information about their lifestyle, physical measures and had blood, urine, and saliva samples collected and stored for future analysis. In this work, the UKB blood biomarkers that are used for the phenotypic predictions consist of 249 Nuclear Magnetic Resonance (NMR) biomarkers from plasma, 31 blood count measures, 30 blood biochemistry measures, 25 infectious disease blood measures, and 3 urine assay measures summing up to a total of 338 initial predictors. In all analyses, age, sex, and statin use were considered as covariates. The inclusion of statin use was necessary due to the significant impact of statins on lipid measures, which are relevant to many predictors. Statin use was derived from the UKB field 20003 surveying prescription drug use. A binary outcome was produced based on the reported use of any HMG-CoA reductase inhibitors including Atorvastatin, Cerivastatin, Fluvastatin, Lovastatin, Mevastatin, Pitavastatin, Pravastatin, Rosuvastatin, Simvastatin, Eptastatin, and Velastatin.

| UKB variables names | UKB category | Counts |
|---|---|---|
| Nuclear Magnetic Resonance (NMR) metabolites | 220 | 249 |
| Blood count | 100081 | 31 |
| Blood biochemistry | 17518 | 30 |
| Infectious diseases | 51428 | 25 |
| Urine assays | 100083 | 3 |

### 5. Supplementary Table T2: Details of UKB Variables

Details of the UKB variable used for training ML methods that contain FieldID, variable name, number of participants, and variable measuring units. More details are available in the UKB online platform.

| FieldID | Field | Participants | Units |
|---|---|---|---|
| 23400 | Total Cholesterol | 275345 | mmol/l |
| 23401 | Total Cholesterol Minus HDL-C | 275345 | mmol/l |
| 23402 | Remnant Cholesterol (Non-HDL, Non-LDL -Cholesterol) | 275344 | mmol/l |

| | | | |
|---|---|---|---|
| 23403 | VLDL Cholesterol | 275344 | mmol/l |
| 23404 | Clinical LDL Cholesterol | 275345 | mmol/l |
| 23405 | LDL Cholesterol | 275344 | mmol/l |
| 23406 | HDL Cholesterol | 275345 | mmol/l |
| 23407 | Total Triglycerides | 275345 | mmol/l |
| 23408 | Triglycerides in VLDL | 275344 | mmol/l |
| 23409 | Triglycerides in LDL | 275344 | mmol/l |
| 23410 | Triglycerides in HDL | 275344 | mmol/l |
| 23411 | Total Phospholipids in Lipoprotein Particles | 275344 | mmol/l |
| 23412 | Phospholipids in VLDL | 275344 | mmol/l |
| 23413 | Phospholipids in LDL | 275344 | mmol/l |
| 23414 | Phospholipids in HDL | 275344 | mmol/l |
| 23415 | Total Esterified Cholesterol | 275344 | mmol/l |
| 23416 | Cholesteryl Esters in VLDL | 275344 | mmol/l |
| 23417 | Cholesteryl Esters in LDL | 275344 | mmol/l |
| 23418 | Cholesteryl Esters in HDL | 275344 | mmol/l |
| 23419 | Total Free Cholesterol | 275344 | mmol/l |
| 23420 | Free Cholesterol in VLDL | 275344 | mmol/l |
| 23421 | Free Cholesterol in LDL | 275344 | mmol/l |
| 23422 | Free Cholesterol in HDL | 275344 | mmol/l |
| 23423 | Total Lipids in Lipoprotein Particles | 275344 | mmol/l |
| 23424 | Total Lipids in VLDL | 275344 | mmol/l |
| 23425 | Total Lipids in LDL | 275344 | mmol/l |

| | | | |
|---|---|---|---|
| 23426 | Total Lipids in HDL | 275344 | mmol/l |
| 23427 | Total Concentration of Lipoprotein Particles | 275344 | mmol/l |
| 23428 | Concentration of VLDL Particles | 275344 | mmol/l |
| 23429 | Concentration of LDL Particles | 275344 | mmol/l |
| 23430 | Concentration of HDL Particles | 275344 | mmol/l |
| 23431 | Average Diameter for VLDL Particles | 275344 | nm |
| 23432 | Average Diameter for LDL Particles | 275344 | nm |
| 23433 | Average Diameter for HDL Particles | 275344 | nm |
| 23434 | Phosphoglycerides | 275137 | mmol/l |
| 23435 | Triglycerides to Phosphoglycerides ratio | 275137 | ratio |
| 23436 | Total Cholines | 275137 | mmol/l |
| 23437 | Phosphatidylcholines | 275137 | mmol/l |
| 23438 | Sphingomyelins | 275131 | mmol/l |
| 23439 | Apolipoprotein B | 275345 | g/l |
| 23440 | Apolipoprotein A1 | 275345 | g/l |
| 23441 | Apolipoprotein B to Apolipoprotein A1 ratio | 275345 | ratio |
| 23442 | Total Fatty Acids | 275137 | mmol/l |
| 23443 | Degree of Unsaturation | 275137 | degree |
| 23444 | Omega-3 Fatty Acids | 275136 | mmol/l |
| 23445 | Omega-6 Fatty Acids | 275136 | mmol/l |
| 23446 | Polyunsaturated Fatty Acids | 275136 | mmol/l |
| 23447 | Monounsaturated Fatty Acids | 275136 | mmol/l |
| 23448 | Saturated Fatty Acids | 275136 | mmol/l |

| | | | |
|---|---|---|---|
| 23449 | Linoleic Acid | 275136 | mmol/l |
| 23450 | Docosahexaenoic Acid | 275136 | mmol/l |
| 23451 | Omega-3 Fatty Acids to Total Fatty Acids percentage | 275136 | percent |
| 23452 | Omega-6 Fatty Acids to Total Fatty Acids percentage | 275136 | percent |
| 23453 | Polyunsaturated Fatty Acids to Total Fatty Acids percentage | 275136 | percent |
| 23454 | Monounsaturated Fatty Acids to Total Fatty Acids percentage | 275136 | percent |
| 23455 | Saturated Fatty Acids to Total Fatty Acids percentage | 275136 | percent |
| 23456 | Linoleic Acid to Total Fatty Acids percentage | 275136 | percent |
| 23457 | Docosahexaenoic Acid to Total Fatty Acids percentage | 275136 | percent |
| 23458 | Polyunsaturated Fatty Acids to Monounsaturated Fatty Acids ratio | 275135 | ratio |
| 23459 | Omega-6 Fatty Acids to Omega-3 Fatty Acids ratio | 275116 | ratio |
| 23460 | Alanine | 275246 | mmol/l |
| 23461 | Glutamine | 274522 | mmol/l |
| 23462 | Glycine | 274973 | mmol/l |
| 23463 | Histidine | 274996 | mmol/l |
| 23464 | Total Concentration of Branched-Chain Amino Acids (Leucine + Isoleucine + Valine) | 275119 | mmol/l |
| 23465 | Isoleucine | 275305 | mmol/l |
| 23466 | Leucine | 275319 | mmol/l |
| 23467 | Valine | 275127 | mmol/l |
| 23468 | Phenylalanine | 275225 | mmol/l |
| 23469 | Tyrosine | 275036 | mmol/l |
| 23470 | Glucose | 274886 | mmol/l |
| 23471 | Lactate | 274854 | mmol/l |

| | | | |
|---|---|---|---|
| 23472 | Pyruvate | 274509 | mmol/l |
| 23473 | Citrate | 275319 | mmol/l |
| 23474 | 3-Hydroxybutyrate | 270488 | mmol/l |
| 23475 | Acetate | 275154 | mmol/l |
| 23476 | Acetoacetate | 275335 | mmol/l |
| 23477 | Acetone | 275343 | mmol/l |
| 23478 | Creatinine | 268823 | mmol/l |
| 23479 | Albumin | 275307 | g/l |
| 23480 | Glycoprotein Acetyls | 275346 | mmol/l |
| 23481 | Concentration of Chylomicrons and Extremely Large VLDL Particles | 275344 | mmol/l |
| 23482 | Total Lipids in Chylomicrons and Extremely Large VLDL | 275344 | mmol/l |
| 23483 | Phospholipids in Chylomicrons and Extremely Large VLDL | 275344 | mmol/l |
| 23484 | Cholesterol in Chylomicrons and Extremely Large VLDL | 275344 | mmol/l |
| 23485 | Cholesteryl Esters in Chylomicrons and Extremely Large VLDL | 275344 | mmol/l |
| 23486 | Free Cholesterol in Chylomicrons and Extremely Large VLDL | 275344 | mmol/l |
| 23487 | Triglycerides in Chylomicrons and Extremely Large VLDL | 275344 | mmol/l |
| 23488 | Concentration of Very Large VLDL Particles | 275344 | mmol/l |
| 23489 | Total Lipids in Very Large VLDL | 275344 | mmol/l |
| 23490 | Phospholipids in Very Large VLDL | 275344 | mmol/l |
| 23491 | Cholesterol in Very Large VLDL | 275344 | mmol/l |
| 23492 | Cholesteryl Esters in Very Large VLDL | 275344 | mmol/l |
| 23493 | Free Cholesterol in Very Large VLDL | 275344 | mmol/l |
| 23494 | Triglycerides in Very Large VLDL | 275344 | mmol/l |

| | | | |
|---|---|---|---|
| 23495 | Concentration of Large VLDL Particles | 275344 | mmol/l |
| 23496 | Total Lipids in Large VLDL | 275344 | mmol/l |
| 23497 | Phospholipids in Large VLDL | 275344 | mmol/l |
| 23498 | Cholesterol in Large VLDL | 275344 | mmol/l |
| 23499 | Cholesteryl Esters in Large VLDL | 275344 | mmol/l |
| 23500 | Free Cholesterol in Large VLDL | 275344 | mmol/l |
| 23501 | Triglycerides in Large VLDL | 275344 | mmol/l |
| 23502 | Concentration of Medium VLDL Particles | 275344 | mmol/l |
| 23503 | Total Lipids in Medium VLDL | 275344 | mmol/l |
| 23504 | Phospholipids in Medium VLDL | 275344 | mmol/l |
| 23505 | Cholesterol in Medium VLDL | 275344 | mmol/l |
| 23506 | Cholesteryl Esters in Medium VLDL | 275344 | mmol/l |
| 23507 | Free Cholesterol in Medium VLDL | 275344 | mmol/l |
| 23508 | Triglycerides in Medium VLDL | 275344 | mmol/l |
| 23509 | Concentration of Small VLDL Particles | 275344 | mmol/l |
| 23510 | Total Lipids in Small VLDL | 275344 | mmol/l |
| 23511 | Phospholipids in Small VLDL | 275344 | mmol/l |
| 23512 | Cholesterol in Small VLDL | 275344 | mmol/l |
| 23513 | Cholesteryl Esters in Small VLDL | 275344 | mmol/l |
| 23514 | Free Cholesterol in Small VLDL | 275344 | mmol/l |
| 23515 | Triglycerides in Small VLDL | 275344 | mmol/l |
| 23516 | Concentration of Very Small VLDL Particles | 275344 | mmol/l |
| 23517 | Total Lipids in Very Small VLDL | 275344 | mmol/l |

| | | | |
|---|---|---|---|
| 23518 | Phospholipids in Very Small VLDL | 275344 | mmol/l |
| 23519 | Cholesterol in Very Small VLDL | 275344 | mmol/l |
| 23520 | Cholesteryl Esters in Very Small VLDL | 275344 | mmol/l |
| 23521 | Free Cholesterol in Very Small VLDL | 275344 | mmol/l |
| 23522 | Triglycerides in Very Small VLDL | 275344 | mmol/l |
| 23523 | Concentration of IDL Particles | 275344 | mmol/l |
| 23524 | Total Lipids in IDL | 275344 | mmol/l |
| 23525 | Phospholipids in IDL | 275344 | mmol/l |
| 23526 | Cholesterol in IDL | 275344 | mmol/l |
| 23527 | Cholesteryl Esters in IDL | 275344 | mmol/l |
| 23528 | Free Cholesterol in IDL | 275344 | mmol/l |
| 23529 | Triglycerides in IDL | 275344 | mmol/l |
| 23530 | Concentration of Large LDL Particles | 275344 | mmol/l |
| 23531 | Total Lipids in Large LDL | 275344 | mmol/l |
| 23532 | Phospholipids in Large LDL | 275344 | mmol/l |
| 23533 | Cholesterol in Large LDL | 275344 | mmol/l |
| 23534 | Cholesteryl Esters in Large LDL | 275344 | mmol/l |
| 23535 | Free Cholesterol in Large LDL | 275344 | mmol/l |
| 23536 | Triglycerides in Large LDL | 275344 | mmol/l |
| 23537 | Concentration of Medium LDL Particles | 275344 | mmol/l |
| 23538 | Total Lipids in Medium LDL | 275344 | mmol/l |
| 23539 | Phospholipids in Medium LDL | 275344 | mmol/l |
| 23540 | Cholesterol in Medium LDL | 275344 | mmol/l |

| | | | |
|---|---|---|---|
| 23541 | Cholesteryl Esters in Medium LDL | 275344 | mmol/l |
| 23542 | Free Cholesterol in Medium LDL | 275344 | mmol/l |
| 23543 | Triglycerides in Medium LDL | 275344 | mmol/l |
| 23544 | Concentration of Small LDL Particles | 275344 | mmol/l |
| 23545 | Total Lipids in Small LDL | 275344 | mmol/l |
| 23546 | Phospholipids in Small LDL | 275344 | mmol/l |
| 23547 | Cholesterol in Small LDL | 275344 | mmol/l |
| 23548 | Cholesteryl Esters in Small LDL | 275344 | mmol/l |
| 23549 | Free Cholesterol in Small LDL | 275344 | mmol/l |
| 23550 | Triglycerides in Small LDL | 275344 | mmol/l |
| 23551 | Concentration of Very Large HDL Particles | 275344 | mmol/l |
| 23552 | Total Lipids in Very Large HDL | 275344 | mmol/l |
| 23553 | Phospholipids in Very Large HDL | 275344 | mmol/l |
| 23554 | Cholesterol in Very Large HDL | 275344 | mmol/l |
| 23555 | Cholesteryl Esters in Very Large HDL | 275344 | mmol/l |
| 23556 | Free Cholesterol in Very Large HDL | 275344 | mmol/l |
| 23557 | Triglycerides in Very Large HDL | 275344 | mmol/l |
| 23558 | Concentration of Large HDL Particles | 275344 | mmol/l |
| 23559 | Total Lipids in Large HDL | 275344 | mmol/l |
| 23560 | Phospholipids in Large HDL | 275344 | mmol/l |
| 23561 | Cholesterol in Large HDL | 275344 | mmol/l |
| 23562 | Cholesteryl Esters in Large HDL | 275344 | mmol/l |
| 23563 | Free Cholesterol in Large HDL | 275344 | mmol/l |

| | | | |
|---|---|---|---|
| 23564 | Triglycerides in Large HDL | 275344 | mmol/l |
| 23565 | Concentration of Medium HDL Particles | 275344 | mmol/l |
| 23566 | Total Lipids in Medium HDL | 275344 | mmol/l |
| 23567 | Phospholipids in Medium HDL | 275344 | mmol/l |
| 23568 | Cholesterol in Medium HDL | 275344 | mmol/l |
| 23569 | Cholesteryl Esters in Medium HDL | 275344 | mmol/l |
| 23570 | Free Cholesterol in Medium HDL | 275344 | mmol/l |
| 23571 | Triglycerides in Medium HDL | 275344 | mmol/l |
| 23572 | Concentration of Small HDL Particles | 275344 | mmol/l |
| 23573 | Total Lipids in Small HDL | 275344 | mmol/l |
| 23574 | Phospholipids in Small HDL | 275344 | mmol/l |
| 23575 | Cholesterol in Small HDL | 275344 | mmol/l |
| 23576 | Cholesteryl Esters in Small HDL | 275344 | mmol/l |
| 23577 | Free Cholesterol in Small HDL | 275344 | mmol/l |
| 23578 | Triglycerides in Small HDL | 275344 | mmol/l |
| 23579 | Phospholipids to Total Lipids in Chylomicrons and Extremely Large VLDL percentage | 264139 | percent |
| 23580 | Cholesterol to Total Lipids in Chylomicrons and Extremely Large VLDL percentage | 264139 | percent |
| 23581 | Cholesteryl Esters to Total Lipids in Chylomicrons and Extremely Large VLDL percentage | 264139 | percent |
| 23582 | Free Cholesterol to Total Lipids in Chylomicrons and Extremely Large VLDL percentage | 264139 | percent |
| 23583 | Triglycerides to Total Lipids in Chylomicrons and Extremely Large VLDL percentage | 264139 | percent |
| 23584 | Phospholipids to Total Lipids in Very Large VLDL percentage | 272820 | percent |
| 23585 | Cholesterol to Total Lipids in Very Large VLDL percentage | 272820 | percent |

| | | | |
|---|---|---|---|
| 23586 | Cholesteryl Esters to Total Lipids in Very Large VLDL percentage | 272820 | percent |
| 23587 | Free Cholesterol to Total Lipids in Very Large VLDL percentage | 272820 | percent |
| 23588 | Triglycerides to Total Lipids in Very Large VLDL percentage | 272820 | percent |
| 23589 | Phospholipids to Total Lipids in Large VLDL percentage | 275312 | percent |
| 23590 | Cholesterol to Total Lipids in Large VLDL percentage | 275312 | percent |
| 23591 | Cholesteryl Esters to Total Lipids in Large VLDL percentage | 275312 | percent |
| 23592 | Free Cholesterol to Total Lipids in Large VLDL percentage | 275312 | percent |
| 23593 | Triglycerides to Total Lipids in Large VLDL percentage | 275312 | percent |
| 23594 | Phospholipids to Total Lipids in Medium VLDL percentage | 275344 | percent |
| 23595 | Cholesterol to Total Lipids in Medium VLDL percentage | 275344 | percent |
| 23596 | Cholesteryl Esters to Total Lipids in Medium VLDL percentage | 275344 | percent |
| 23597 | Free Cholesterol to Total Lipids in Medium VLDL percentage | 275344 | percent |
| 23598 | Triglycerides to Total Lipids in Medium VLDL percentage | 275344 | percent |
| 23599 | Phospholipids to Total Lipids in Small VLDL percentage | 275344 | percent |
| 23600 | Cholesterol to Total Lipids in Small VLDL percentage | 275344 | percent |
| 23601 | Cholesteryl Esters to Total Lipids in Small VLDL percentage | 275344 | percent |
| 23602 | Free Cholesterol to Total Lipids in Small VLDL percentage | 275344 | percent |
| 23603 | Triglycerides to Total Lipids in Small VLDL percentage | 275344 | percent |
| 23604 | Phospholipids to Total Lipids in Very Small VLDL percentage | 275344 | percent |
| 23605 | Cholesterol to Total Lipids in Very Small VLDL percentage | 275344 | percent |
| 23606 | Cholesteryl Esters to Total Lipids in Very Small VLDL percentage | 275344 | percent |
| 23607 | Free Cholesterol to Total Lipids in Very Small VLDL percentage | 275344 | percent |
| 23608 | Triglycerides to Total Lipids in Very Small VLDL percentage | 275344 | percent |

| | | | |
|---|---|---|---|
| 23609 | Phospholipids to Total Lipids in IDL percentage | 275344 | percent |
| 23610 | Cholesterol to Total Lipids in IDL percentage | 275344 | percent |
| 23611 | Cholesteryl Esters to Total Lipids in IDL percentage | 275344 | percent |
| 23612 | Free Cholesterol to Total Lipids in IDL percentage | 275344 | percent |
| 23613 | Triglycerides to Total Lipids in IDL percentage | 275344 | percent |
| 23614 | Phospholipids to Total Lipids in Large LDL percentage | 275344 | percent |
| 23615 | Cholesterol to Total Lipids in Large LDL percentage | 275344 | percent |
| 23616 | Cholesteryl Esters to Total Lipids in Large LDL percentage | 275344 | percent |
| 23617 | Free Cholesterol to Total Lipids in Large LDL percentage | 275344 | percent |
| 23618 | Triglycerides to Total Lipids in Large LDL percentage | 275344 | percent |
| 23619 | Phospholipids to Total Lipids in Medium LDL percentage | 275344 | percent |
| 23620 | Cholesterol to Total Lipids in Medium LDL percentage | 275344 | percent |
| 23621 | Cholesteryl Esters to Total Lipids in Medium LDL percentage | 275344 | percent |
| 23622 | Free Cholesterol to Total Lipids in Medium LDL percentage | 275344 | percent |
| 23623 | Triglycerides to Total Lipids in Medium LDL percentage | 275344 | percent |
| 23624 | Phospholipids to Total Lipids in Small LDL percentage | 275344 | percent |
| 23625 | Cholesterol to Total Lipids in Small LDL percentage | 275344 | percent |
| 23626 | Cholesteryl Esters to Total Lipids in Small LDL percentage | 275344 | percent |
| 23627 | Free Cholesterol to Total Lipids in Small LDL percentage | 275344 | percent |
| 23628 | Triglycerides to Total Lipids in Small LDL percentage | 275344 | percent |
| 23629 | Phospholipids to Total Lipids in Very Large HDL percentage | 275238 | percent |
| 23630 | Cholesterol to Total Lipids in Very Large HDL percentage | 275238 | percent |
| 23631 | Cholesteryl Esters to Total Lipids in Very Large HDL percentage | 275238 | percent |

| | | | |
|---|---|---|---|
| 23632 | Free Cholesterol to Total Lipids in Very Large HDL percentage | 275238 | percent |
| 23633 | Triglycerides to Total Lipids in Very Large HDL percentage | 275238 | percent |
| 23634 | Phospholipids to Total Lipids in Large HDL percentage | 275344 | percent |
| 23635 | Cholesterol to Total Lipids in Large HDL percentage | 275344 | percent |
| 23636 | Cholesteryl Esters to Total Lipids in Large HDL percentage | 275344 | percent |
| 23637 | Free Cholesterol to Total Lipids in Large HDL percentage | 275344 | percent |
| 23638 | Triglycerides to Total Lipids in Large HDL percentage | 275344 | percent |
| 23639 | Phospholipids to Total Lipids in Medium HDL percentage | 275344 | percent |
| 23640 | Cholesterol to Total Lipids in Medium HDL percentage | 275344 | percent |
| 23641 | Cholesteryl Esters to Total Lipids in Medium HDL percentage | 275344 | percent |
| 23642 | Free Cholesterol to Total Lipids in Medium HDL percentage | 275344 | percent |
| 23643 | Triglycerides to Total Lipids in Medium HDL percentage | 275344 | percent |
| 23644 | Phospholipids to Total Lipids in Small HDL percentage | 275344 | percent |
| 23645 | Cholesterol to Total Lipids in Small HDL percentage | 275344 | percent |
| 23646 | Cholesteryl Esters to Total Lipids in Small HDL percentage | 275344 | percent |
| 23647 | Free Cholesterol to Total Lipids in Small HDL percentage | 275344 | percent |
| 23648 | Triglycerides to Total Lipids in Small HDL percentage | 275344 | percent |
| 30000 | White blood cell (leukocyte) count | 479215 | 10^9 cells/Litre |
| 30010 | Red blood cell (erythrocyte) count | 479220 | 10^12 cells/Litre |
| 30020 | Haemoglobin concentration | 479219 | grams/decilitre |
| 30030 | Haematocrit percentage | 479220 | percent |
| 30040 | Mean corpuscular volume | 479219 | femtolitres |

| | | | |
|---|---|---|---|
| 30050 | Mean corpuscular haemoglobin | 479216 | picograms |
| 30060 | Mean corpuscular haemoglobin concentration | 479213 | grams/decilitre |
| 30070 | Red blood cell (erythrocyte) distribution width | 479219 | percent |
| 30080 | Platelet count | 479216 | 10^9 cells/Litre |
| 30090 | Platelet crit | 479002 | percent |
| 30100 | Mean platelet (thrombocyte) volume | 479211 | femtolitres |
| 30110 | Platelet distribution width | 479001 | percent |
| 30120 | Lymphocyte count | 478352 | 10^9 cells/Litre |
| 30130 | Monocyte count | 478352 | 10^9 cells/Litre |
| 30140 | Neutrophill count | 478352 | 10^9 cells/Litre |
| 30150 | Eosinophill count | 478352 | 10^9 cells/Litre |
| 30160 | Basophill count | 478352 | 10^9 cells/Litre |
| 30170 | Nucleated red blood cell count | 478126 | 10^9 cells/Litre |
| 30180 | Lymphocyte percentage | 478357 | percent |
| 30190 | Monocyte percentage | 478357 | percent |
| 30200 | Neutrophill percentage | 478357 | percent |
| 30210 | Eosinophill percentage | 478357 | percent |
| 30220 | Basophill percentage | 478357 | percent |
| 30230 | Nucleated red blood cell percentage | 478122 | percent |
| 30240 | Reticulocyte percentage | 471169 | percent |
| 30250 | Reticulocyte count | 471169 | 10^12 cells/Litre |

| | | | |
|---|---|---|---|
| 30260 | Mean reticulocyte volume | 471168 | femtolitres |
| 30270 | Mean sphered cell volume | 470909 | femtolitres |
| 30280 | Immature reticulocyte fraction | 470907 | ratio |
| 30290 | High light scatter reticulocyte percentage | 470909 | percent |
| 30300 | High light scatter reticulocyte count | 470908 | 10^12 cells/Litre |
| 30600 | Albumin | 432221 | g/L |
| 30610 | Alkaline phosphatase | 470729 | U/L |
| 30620 | Alanine aminotransferase | 470528 | U/L |
| 30630 | Apolipoprotein A | 429675 | g/L |
| 30640 | Apolipoprotein B | 468384 | g/L |
| 30650 | Aspartate aminotransferase | 468980 | U/L |
| 30660 | Direct bilirubin | 400913 | umol/L |
| 30670 | Urea | 470408 | mmol/L |
| 30680 | Calcium | 432073 | mmol/L |
| 30690 | Cholesterol | 470716 | mmol/L |
| 30700 | Creatinine | 470493 | umol/L |
| 30710 | C-reactive protein | 469727 | mg/L |
| 30720 | Cystatin C | 470687 | mg/L |
| 30730 | Gamma glutamyltransferase | 470474 | U/L |
| 30740 | Glucose | 431727 | mmol/L |
| 30750 | Glycated haemoglobin (HbA1c) | 467782 | mmol/mol |
| 30760 | HDL cholesterol | 432018 | mmol/L |
| 30770 | IGF-1 | 468257 | nmol/L |

| | | | |
|---|---|---|---|
| 30780 | LDL direct | 469878 | mmol/L |
| 30790 | Lipoprotein A | 377555 | nmol/L |
| 30800 | Oestradiol | 77678 | pmol/L |
| 30810 | Phosphate | 431408 | mmol/L |
| 30820 | Rheumatoid factor | 41979 | IU/ml |
| 30830 | SHBG | 428060 | nmol/L |
| 30840 | Total bilirubin | 468757 | umol/L |
| 30850 | Testosterone | 426940 | nmol/L |
| 30860 | Total protein | 431755 | g/L |
| 30870 | Triglycerides | 470346 | mmol/L |
| 30880 | Urate | 470167 | umol/L |
| 30890 | Vitamin D | 449830 | nmol/L |
| 23050 | HSV-1 seropositivity for Herpes Simplex virus-1 | 9689 | |
| 23051 | HSV-2 seropositivity for Herpes Simplex virus-2 | 9689 | |
| 23052 | VZV seropositivity for Varicella Zoster Virus | 9689 | |
| 23053 | EBV seropositivity for Epstein-Barr Virus | 9689 | |
| 23054 | CMV seropositivity for Human Cytomegalovirus | 9689 | |
| 23055 | HHV-6 overall seropositivity for Human Herpesvirus-6 | 9689 | |
| 23056 | HHV-6A seropositivity for Human Herpesvirus-6 | 9689 | |
| 23057 | HHV-6B seropositivity for Human Herpesvirus-6 | 9689 | |
| 23058 | HHV-7 seropositivity for Human Herpesvirus-7 | 9689 | |
| 23059 | KSHV seropositivity for Kaposi's Sarcoma-Associated Herpesvirus | 9689 | |
| 23060 | HBV seropositivity for Hepatitis B Virus | 9689 | |

| | | | |
|---|---|---|---|
| 23061 | HCV seropositivity for Hepatitis C Virus | 9689 | |
| 23062 | T. gondii seropositivity for Toxoplasma gondii | 9689 | |
| 23063 | HTLV-1 seropositivity for Human T-Lymphotropic Virus 1 | 9689 | |
| 23064 | HIV-1 seropositivity for Human Immunodeficiency Virus | 9689 | |
| 23065 | BKV seropositivity for Human Polyomavirus BKV | 9689 | |
| 23066 | JCV seropositivity for Human Polyomavirus JCV | 9689 | |
| 23067 | MCV seropositivity for Merkel Cell Polyomavirus | 9689 | |
| 23068 | HPV 16 Definition I seropositivity for Human Papillomavirus type-16 | 9689 | |
| 23069 | HPV 18 seropositivity for Human Papillomavirus type-18 | 9689 | |
| 23070 | C. trachomatis Definition I seropositivity for Chlamydia trachomatis | 9689 | |
| 23071 | C. trachomatis Definition II seropositivity for Chlamydia trachomatis | 7618 | |
| 23073 | H. pylori Definition I seropositivity for Helicobacter pylori | 4897 | |
| 23074 | H. pylori Definition II seropositivity for Helicobacter pylori | 9689 | |
| 23075 | HPV 16 Definition II seropositivity for Human Papillomavirus type-16 | 9689 | |
| 30510 | Creatinine (enzymatic) in urine | 484824 | micromole/L |
| 30520 | Potassium in urine | 483840 | millimole/L |
| 30530 | Sodium in urine | 483810 | millimole/L |

### 6. Supplementary Table T3: Data filtering and splitting

Data filtering involves multiple tasks such as labeling, handling missing values, encoding categorical variables, scaling, and normalizing numerical variables. The UK Biobank is one of the largest and most comprehensive population-based biobanks in the world, collecting a wide range of health-related data from hundreds of thousands of participants. Given the volume and complexity of UKB data, careful data treatment is required to guarantee satisfactory results. Characterizing patterns of missingness, and standardizing data to a common format and units,

especially when data come from diverse sources, are all essential before feeding features into the ML pipeline to achieve meaningful results.
The starting sample size consists of 359,980 independent European UKB individuals. The final sample size and predictor variables were determined for each outcome separately using a four-step filtering process. First, subjects with the missing outcome of interest were dropped. Second, subjects missing NMR metabolite data were dropped, as this assay represents the primary predictors of interest. Third, predictor variables missing in 10% or more of the remaining subjects were dropped. Finally, any subjects with missingness in the remaining predictors were dropped.

Data splitting divides the dataset into multiple subsets for training, validation, and testing purposes. First, the entire dataset is split into a test and a training set, typically along a 20/80 (test/train) split. The training set is used to estimate, assess, and hyper-tune the ML model's parameters, while the test set is completely withheld to validate the final ML model. Additional data splitting within the training set is necessary to estimate the optimal ML model for each implementation called a validation set. Within the training set, data splitting is accomplished by K-Fold Cross-Validation (CV,) a widely used technique in ML and statistical modeling for assessing predictive model performance and generalizability. The typical value for K-Fold Cross-Validation is 5. The sample sizes for all data filtering and splitting stages are shown in the table below.

| **Phenotype** | **Sample Counts** | **Sample Counts (remove missing for primary outcome)** | **Sample Counts (Remove missing with metabolite critical)** | **Sample Counts (Remove missing variables with excessive missingness of more than 10%)** | **Sample Counts (Remove any missing values)** | **Training** | **Validation** | **Testing** |
|---|---|---|---|---|---|---|---|---|
| height_50 | 359908 | 359118 | 86381 | 86381 | 63018 | 40263 | 10066 | 12689 |
| bmi_21001 | 359908 | 358720 | 86282 | 86282 | 62955 | 40220 | 10055 | 12680 |
| alcDPW_all_base | 359908 | 359564 | 86468 | 86468 | 63058 | 40290 | 10072 | 12696 |
| BodFatPer_23099 | 359908 | 353397 | 84995 | 84995 | 62030 | 39617 | 9904 | 12509 |
| MDD | 359908 | 117560 | 28222 | 28222 | 20578 | 13191 | 3298 | 4089 |

7. **Supplementary Table T4: Adjusted $R^2$ for the predicted height, BMI, DPW, BF%, and MDDsx**.
The training process is organized into three steps in order of increasing model complexity and prediction accuracy. First, a linear regression model is trained using only age, sex, and statin use

as predictors. Second, a linear regression model is trained on age, sex, and statin use, in addition to four blood biochemistry assay metrics of liver function: alkaline phosphatase (AlkPhos), alanine aminotransferase (ALT), aspartate aminotransferase (AST), and gamma-glutamyltransferase (GGT). The third and final models were trained using five ML frameworks to include all blood-based biomarkers, in addition to age, sex, and statin use.

| Phenotype | Covariates Only | Covariates & Liver Enzymes Only | LASSO | RIDGE | MBOOST | GBM | XGBOOST |
|---|---|---|---|---|---|---|---|
| height | 0.525 | 0.528 | 0.545 | 0.54 | 0.542 | 0.548 | 0.543 |
| BMI | 0.009 | 0.099 | 0.455 | 0.445 | 0.426 | 0.486 | 0.496 |
| DPW | 0.071 | 0.105 | 0.333 | 0.323 | 0.304 | 0.354 | 0.356 |
| BF% | 0.476 | 0.531 | 0.709 | 0.693 | 0.69 | 0.725 | 0.727 |
| MDDsx | 0.069 | 0.069 | 0.002 | 0.002 | 0.001 | 0.002 | 0.002 |

8. **Supplementary Table T5: MAE for the predicted height, BMI, DPW, BF%, and MDDsx.** Estimate MAE for three ML models. First, a linear regression model is trained using only age, sex, and statin use as predictors. Second, a linear regression model is trained on age, sex, and statin use, in addition to four blood biochemistry assay metrics of liver function: alkaline phosphatase (AlkPhos), alanine aminotransferase (ALT), aspartate aminotransferase (AST), and gamma-glutamyltransferase (GGT). The third and final models were trained using five ML frameworks to include all blood-based biomarkers, in addition to age, sex, and statin use.

| Phenotype | Covariates Only | Covariates & Liver Enzymes Only | LASSO | RIDGE | MBOOST | GBM | XGBOOST |
|---|---|---|---|---|---|---|---|
| height | 5.053 | 5.034 | 4.897 | 4.922 | 4.911 | 4.879 | 4.903 |
| BMI | 3.511 | 3.298 | 2.557 | 2.574 | 2.606 | 2.483 | 2.451 |
| DPW | 6.511 | 6.363 | 5.403 | 5.435 | 5.51 | 5.265 | 5.214 |
| BF% | 4.879 | 4.573 | 3.564 | 3.657 | 3.665 | 3.481 | 3.468 |
| MDDsx | 2.582 | 2.581 | 2.568 | 2.568 | 2.575 | 2.584 | 2.59 |

9. **Supplementary Table T6: Estimated heritability** LDSC estimated heritabilities for observed and predicted DPW including labeling of seven sets, sample size, intercept, and heritability with standard error.

| Outcome | Sample size | intercept | $h^2$(SE) |
|---|---|---|---|
| Unfiltered Set | 359564 | 1.004 | 0.0643 (0.0031) |

| | | | |
|---|---|---|---|
| Filtered Set | 63058 | 0.9898 | 0.0748 (0.0095) |
| Predicted DPW (LASSO) | 63058 | 0.9843 | 0.1428 (0.014) |
| Predicted DPW (RIDGE) | 63058 | 0.9822 | 0.1457 (0.0146) |
| Predicted DPW (MBOOST) | 63058 | 0.9958 | 0.1539 (0.0165) |
| Predicted DPW (GBM) | 63058 | 0.9903 | 0.1234 (0.0121) |
| Predicted DPW (XGBoost) | 63058 | 0.9958 | 0.0776 (0.0091) |

### 10. Supplementary Table T7: Estimated genetic correlations

LDSC estimated genetic correlations for observed and predicted DPW including labeling of 21 sets, genetic correlation with standard error, and p-value. The sets are generated by a pairwise combination of sets defined in supplementary Table T6.

| **Outcome** | **$r_G$(SE)** | **p-value** |
|---|---|---|
| Unfiltered Set vs Filtered Set | 0.9434 (0.0404) | 2.31E-120 |
| Unfiltered Set vs LASSO | 0.5338 (0.0381) | 1.11E-44 |
| Unfiltered Set vs RIDGE | 0.5212 (0.0381) | 1.58E-42 |
| Unfiltered Set vs MBOOST | 0.4911 (0.0372) | 8.80E-40 |
| Unfiltered Set vs GBM | 0.5709 (0.0353) | 1.04E-58 |
| Unfiltered Set vs XGBOOST | 0.8774 (0.0405) | 2.87E-104 |
| Filtered Set vs LASSO | 0.5037 (0.0471) | 1.07E-26 |
| Filtered Set vs RIDGE | 0.4931 (0.0461) | 1.18E-26 |
| Filtered Set vs MBOOST | 0.4535 (0.0471) | 6.06E-22 |
| Filtered Set vs GBM | 0.5063 (0.0466) | 1.57E-27 |
| Filtered Set vs XGBOOST | 0.8191 (0.0246) | 2.00E-242 |
| LASSO vs RIDGE | 0.9981 (0.0007) | 0.00E+00 |
| LASSO vs MBOOST | 0.9797 (0.0043) | 0.00E+00 |
| LASSO vs GBM | 0.9721 (0.0071) | 0.00E+00 |
| LASSO vs XGBOOST | 0.8983 (0.0214) | 0.00E+00 |
| RIDGE vs MBOOST | 0.9875 (0.0031) | 0.00E+00 |
| RIDGE vs GBM | 0.9775 (0.006) | 0.00E+00 |
| RIDGE vs XGBOOST | 0.8916 (0.0206) | 0.00E+00 |

| | | |
|---|---|---|
| MBOOST vs GBM | 0.9801 (0.0061) | 0.00E+00 |
| MBOOST vs XGBOOST | 0.8635 (0.0226) | 0.00E+00 |
| GBM vs XGBOOST | 0.9039 (0.0163) | 0.00E+00 |

### 11. Supplementary Table T8: Confusion matrices for DPW>8

Confusion matrices for predicted DPW>8 for females across five ML models.

| ML model | | |
|---|---|---|
| | Reference | |
| Prediction | 0 (DPW<8) | 1 (DPW>8) |
| 0 | TN | FN |
| 1 | FP | TP |

| MBOOST | | |
|---|---|---|
| | Reference | |
| Prediction | 0 | 1 |
| 0 | 19663 | 3442 |
| 1 | 4756 | 4722 |

| LASSO | | |
|---|---|---|
| | Reference | |
| Prediction | 0 | 1 |
| 0 | 20724 | 3744 |
| 1 | 3695 | 4420 |

| GBM | | |
|---|---|---|
| | Reference | |
| Prediction | 0 | 1 |
| 0 | 20840 | 3704 |
| 1 | 3579 | 4460 |

| RIDGE | | |
|---|---|---|
| | Reference | |
| Prediction | 0 | 1 |
| 0 | 20586 | 3738 |
| 1 | 3833 | 4426 |

| XGBOOST | | |
|---|---|---|
| | Reference | |
| Prediction | 0 | 1 |
| 0 | 22713 | 2271 |
| 1 | 1706 | 5893 |

### 12. Supplementary Table T9: Confusion matrices for DPW>15

Confusion matrices for predicted DPW>15 for males across five ML models.

| ML model | | |
|---|---|---|
| | Reference | |
| Prediction | 0 (DPW<15) | 1 (DPW>15) |
| 0 | TN | FN |

| MBOOST | | |
|---|---|---|
| | Reference | |
| Prediction | 0 | 1 |
| 0 | 20766 | 4351 |

| | | | | | | |
|---|---|---|---|---|---|---|
| 1 | FP | TP | | 1 | 1933 | 3425 |
| **LASSO** | | | | **GBM** | | |
| | Reference | | | | Reference | |
| Prediction | 0 | 1 | | Prediction | 0 | 1 |
| 0 | 20940 | 4313 | | 0 | 20417 | 3795 |
| 1 | 1759 | 3463 | | 1 | 2282 | 3981 |
| **RIDGE** | | | | **XGBOOST** | | |
| | Reference | | | | Reference | |
| Prediction | 0 | 1 | | Prediction | 0 | 1 |
| 0 | 21019 | 4431 | | 0 | 21410 | 2063 |
| 1 | 1680 | 3345 | | 1 | 1289 | 5713 |

### 13. Supplementary Table T10: Evaluation of ML metrics for DPW>8

ML Metrics evaluation including Accuracy, F1 score, Sensitivity (Recall), Specificity, Positive Predictive Value (PPV) (Precision), and Negative Predictive Value (NPV) for females and predicted DPW>8 across five ML models.

| ML Model | Accuracy | F1 score | Sensitivity (Recall) | Specificity | PPV (Precision) | NPV |
|---|---|---|---|---|---|---|
| LASSO | 77.17% | 54.30% | 54.14% | 84.87% | 54.47% | 84.70% |
| RIDGE | 76.76% | 53.90% | 54.21% | 84.30% | 53.59% | 84.63% |
| GBM | 77.65% | 55.05% | 54.63% | 85.34% | 55.48% | 84.91% |
| MBOOST | 74.84% | 53.53% | 57.84% | 80.52% | 49.82% | 85.10% |
| XGBOOST | 87.79% | 74.77% | 72.18% | 93.01% | 77.55% | 90.91% |

### 14. Supplementary Table T11: Evaluation of ML metrics for DPW>15

ML Metrics evaluation including Accuracy, Recall, Precision, F1 score, Sensitivity (Recall), Specificity, Positive Predictive Value (PPV) (Precision), and Negative Predictive Value (NPV) for males and predicted DPW>15 across five ML models.

| ML Model | Accuracy | F1 score | Sensitivity (Recall) | Specificity | PPV (Precision) | NPV |
|---|---|---|---|---|---|---|
| LASSO | 80.08% | 53.29% | 44.53% | 92.25% | 66.32% | 82.92% |
| RIDGE | 79.95% | 52.26% | 43.02% | 92.60% | 66.57% | 82.59% |
| GBM | 80.06% | 56.71% | 51.20% | 89.95% | 63.56% | 84.33% |
| MBOOST | 79.38% | 52.15% | 44.05% | 91.48% | 63.92% | 82.68% |
| XGBOOST | 89.00% | 77.32% | 73.47% | 94.32% | 81.59% | 91.21% |

### 15. Supplementary Table T12: A full list of all SHAP values of the blood-based biomarkers used to train ML models.

A full list of SHAP values for all the variables ranked in descending order from the high SHAP score to the lowest score. We used the R package "SHAPforxgboost" and the XGBoost model to generate these values.

| Rank | Variable Name | Mean SHAP Score |
|---|---|---|
| 1 | sex | 1.90580067 |
| 2 | Gamma glutamyltransferase | 0.93174673 |
| 3 | Citrate | 0.8381188 |
| 4 | Phospholipids in Medium HDL | 0.76084602 |
| 5 | Urate | 0.69966289 |
| 6 | Saturated Fatty Acids to Total Fatty Acids percentage | 0.51431696 |
| 7 | Mean corpuscular volume | 0.4894054 |
| 8 | 3-Hydroxybutyrate | 0.46767473 |
| 9 | Mean corpuscular haemoglobin | 0.44755549 |
| 10 | Linoleic Acid to Total Fatty Acids percentage | 0.38425705 |
| 11 | Cystatin C | 0.36854069 |
| 12 | Potassium in urine | 0.36183415 |
| 13 | Valine | 0.35656856 |
| 14 | Tyrosine | 0.3428176 |
| 15 | Alkaline phosphatase | 0.31380739 |
| 16 | Total Lipids in Medium HDL | 0.25548201 |
| 17 | Docosahexaenoic Acid to Total Fatty Acids percentage | 0.23466496 |
| 18 | IGF-1 | 0.21105307 |
| 19 | Cholesteryl Esters in Medium HDL | 0.21100754 |

| | | |
|---|---|---|
| 20 | Vitamin D | 0.20769218 |
| 21 | Acetone | 0.19990328 |
| 22 | Free Cholesterol to Total Lipids in IDL percentage | 0.19797213 |
| 23 | Total bilirubin | 0.19610656 |
| 24 | Urea | 0.18712675 |
| 25 | Cholesteryl Esters in Small HDL | 0.17942036 |
| 26 | age | 0.17227394 |
| 27 | Apolipoprotein B | 0.17193944 |
| 28 | Concentration of HDL Particles | 0.1699211 |
| 29 | Glycated haemoglobin (HbA1c) | 0.15673972 |
| 30 | Degree of Unsaturation | 0.15030846 |
| 31 | Monocyte percentage | 0.147083 |
| 32 | Alanine aminotransferase | 0.1398917 |
| 33 | Glucose | 0.13947025 |
| 34 | Phospholipids in Small HDL | 0.13555178 |
| 35 | Mean sphered cell volume | 0.13229389 |
| 36 | Cholesterol | 0.13048113 |
| 37 | C-reactive protein | 0.12136589 |
| 38 | Docosahexaenoic Acid | 0.11628656 |
| 39 | Average Diameter for LDL Particles | 0.11268294 |
| 40 | Lymphocyte count | 0.10978852 |
| 41 | Glycine | 0.10929162 |
| 42 | Sodium in urine | 0.1091487 |
| 43 | Glycoprotein Acetyls | 0.10733032 |
| 44 | Lactate | 0.10681865 |
| 45 | Eosinophill percentage | 0.10421841 |
| 46 | Phospholipids to Total Lipids in Very Large VLDL percentage | 0.10022677 |
| 47 | Acetate | 0.09811672 |
| 48 | Leucine | 0.09515205 |

| | | |
|---|---|---|
| 49 | Free Cholesterol to Total Lipids in Very Large VLDL percentage | 0.09482705 |
| 50 | Phospholipids to Total Lipids in Large HDL percentage | 0.0928367 |
| 51 | Reticulocyte percentage | 0.09080339 |
| 52 | Mean reticulocyte volume | 0.08867555 |
| 53 | Acetoacetate | 0.08844978 |
| 54 | Phospholipids to Total Lipids in Very Small VLDL percentage | 0.08657538 |
| 55 | Platelet distribution width | 0.08650369 |
| 56 | Phenylalanine | 0.08443968 |
| 57 | Cholesterol in Small HDL | 0.08414465 |
| 58 | any_statin | 0.08413941 |
| 59 | Aspartate aminotransferase | 0.08395956 |
| 60 | Triglycerides in Large HDL | 0.08337213 |
| 61 | Phospholipids to Total Lipids in Large VLDL percentage | 0.08294843 |
| 62 | Linoleic Acid | 0.08280047 |
| 63 | Free Cholesterol to Total Lipids in Small HDL percentage | 0.08118407 |
| 64 | High light scatter reticulocyte percentage | 0.07806637 |
| 65 | Cholesteryl Esters in HDL | 0.07763069 |
| 66 | Triglycerides | 0.07707651 |
| 67 | Isoleucine | 0.07659416 |
| 68 | Triglycerides to Total Lipids in Large HDL percentage | 0.07639966 |
| 69 | Triglycerides to Total Lipids in Medium HDL percentage | 0.07238008 |
| 70 | Phospholipids to Total Lipids in Small HDL percentage | 0.0714558 |
| 71 | Monounsaturated Fatty Acids to Total Fatty Acids percentage | 0.0685287 |
| 72 | Cholesteryl Esters to Total Lipids in Chylomicrons and Extremely Large VLDL percentage | 0.0678127 |
| 73 | Lymphocyte percentage | 0.06510694 |
| 74 | Pyruvate | 0.06477977 |
| 75 | Alanine | 0.06389324 |
| 76 | Phospholipids to Total Lipids in Small LDL percentage | 0.06114409 |
| 77 | Mean platelet (thrombocyte) volume | 0.06057632 |

| | | |
|---|---|---|
| 78 | Polyunsaturated Fatty Acids to Total Fatty Acids percentage | 0.0604847 |
| 79 | White blood cell (leukocyte) count | 0.0593689 |
| 80 | Platelet crit | 0.05838319 |
| 81 | Cholesteryl Esters to Total Lipids in Very Large HDL percentage | 0.05740309 |
| 82 | Omega-6 Fatty Acids to Total Fatty Acids percentage | 0.05636871 |
| 83 | Glutamine | 0.0546221 |
| 84 | Red blood cell (erythrocyte) count | 0.05314761 |
| 85 | Polyunsaturated Fatty Acids to Monounsaturated Fatty Acids ratio | 0.04955637 |
| 86 | Monocyte count | 0.04833591 |
| 87 | Creatinine (enzymatic) in urine | 0.04691863 |
| 88 | Creatinine | 0.04681481 |
| 89 | Platelet count | 0.04548887 |
| 90 | Cholesteryl Esters to Total Lipids in Medium LDL percentage | 0.04487482 |
| 91 | Red blood cell (erythrocyte) distribution width | 0.04452361 |
| 92 | Cholesterol in Medium HDL | 0.04449789 |
| 93 | Apolipoprotein B to Apolipoprotein A1 ratio | 0.0444511 |
| 94 | Cholesteryl Esters to Total Lipids in Large HDL percentage | 0.04397287 |
| 95 | Immature reticulocyte fraction | 0.04310637 |
| 96 | Triglycerides in Very Large HDL | 0.04308162 |
| 97 | Total Lipids in Small HDL | 0.04262021 |
| 98 | Cholesterol to Total Lipids in Medium LDL percentage | 0.04248892 |
| 99 | Free Cholesterol to Total Lipids in Very Small VLDL percentage | 0.04080766 |
| 100 | Albumin | 0.04043315 |
| 101 | Cholesteryl Esters to Total Lipids in Large LDL percentage | 0.039086 |
| 102 | Haemoglobin concentration | 0.03814921 |
| 103 | Free Cholesterol to Total Lipids in Chylomicrons and Extremely Large VLDL percentage | 0.03769953 |
| 104 | Cholesterol to Total Lipids in Large HDL percentage | 0.03696371 |
| 105 | Mean corpuscular haemoglobin concentration | 0.03690402 |
| 106 | Creatinine | 0.0366985 |

| | | |
|---|---|---|
| 107 | Histidine | 0.03630687 |
| 108 | Omega-6 Fatty Acids | 0.03545848 |
| 109 | Phosphatidylcholines | 0.03539046 |
| 110 | Free Cholesterol to Total Lipids in Large VLDL percentage | 0.03490089 |
| 111 | Haematocrit percentage | 0.03449607 |
| 112 | Total Concentration of Lipoprotein Particles | 0.0337928 |
| 113 | Cholesterol to Total Lipids in Small LDL percentage | 0.03369107 |
| 114 | Concentration of Small VLDL Particles | 0.03329928 |
| 115 | Triglycerides in Medium HDL | 0.03307925 |
| 116 | Phospholipids in HDL | 0.03301658 |
| 117 | Neutrophill count | 0.03251818 |
| 118 | HDL Cholesterol | 0.03205886 |
| 119 | Basophill percentage | 0.0311865 |
| 120 | Phospholipids to Total Lipids in Medium VLDL percentage | 0.03113048 |
| 121 | Reticulocyte count | 0.0310238 |
| 122 | Concentration of Small HDL Particles | 0.03097428 |
| 123 | Cholesteryl Esters to Total Lipids in Very Large VLDL percentage | 0.03089629 |
| 124 | Total Concentration of Branched-Chain Amino Acids (Leucine + Isoleucine + Valine) | 0.03037616 |
| 125 | Concentration of Medium HDL Particles | 0.02869759 |
| 126 | Apolipoprotein A1 | 0.02835709 |
| 127 | Phospholipids to Total Lipids in IDL percentage | 0.02780802 |
| 128 | Neutrophill percentage | 0.02749975 |
| 129 | Omega-6 Fatty Acids to Omega-3 Fatty Acids ratio | 0.02718001 |
| 130 | Cholesterol to Total Lipids in Chylomicrons and Extremely Large VLDL percentage | 0.02717306 |
| 131 | Sphingomyelins | 0.02587307 |
| 132 | Concentration of Small LDL Particles | 0.02536512 |
| 133 | Phospholipids to Total Lipids in Medium LDL percentage | 0.02513571 |
| 134 | Cholesterol to Total Lipids in Medium HDL percentage | 0.02477918 |

| | | |
|---|---|---|
| 135 | Triglycerides to Total Lipids in Small HDL percentage | 0.02464073 |
| 136 | Cholesteryl Esters to Total Lipids in IDL percentage | 0.02456641 |
| 137 | Free Cholesterol to Total Lipids in Very Large HDL percentage | 0.02436091 |
| 138 | Omega-3 Fatty Acids to Total Fatty Acids percentage | 0.02416507 |
| 139 | Free Cholesterol in Very Large HDL | 0.0239977 |
| 140 | Triglycerides to Phosphoglycerides ratio | 0.02379634 |
| 141 | Cholesterol to Total Lipids in Very Large HDL percentage | 0.02378406 |
| 142 | Eosinophill count | 0.02344085 |
| 143 | Total Lipids in Medium VLDL | 0.02335246 |
| 144 | Polyunsaturated Fatty Acids | 0.02329086 |
| 145 | Total Lipids in HDL | 0.02210996 |
| 146 | Cholesteryl Esters to Total Lipids in Medium HDL percentage | 0.02165424 |
| 147 | Phospholipids to Total Lipids in Large LDL percentage | 0.02162372 |
| 148 | Phospholipids to Total Lipids in Medium HDL percentage | 0.02080332 |
| 149 | Cholesterol to Total Lipids in Large LDL percentage | 0.01989894 |
| 150 | Cholesteryl Esters to Total Lipids in Small VLDL percentage | 0.01961839 |
| 151 | Cholesterol to Total Lipids in IDL percentage | 0.01946205 |
| 152 | Total Lipids in Very Large HDL | 0.01912633 |
| 153 | Free Cholesterol to Total Lipids in Medium LDL percentage | 0.0188579 |
| 154 | Free Cholesterol to Total Lipids in Large HDL percentage | 0.01881123 |
| 155 | Concentration of Very Large HDL Particles | 0.01874137 |
| 156 | Triglycerides in Large LDL | 0.01852025 |
| 157 | Cholesterol to Total Lipids in Small VLDL percentage | 0.01851565 |
| 158 | Cholesteryl Esters to Total Lipids in Small LDL percentage | 0.01845256 |
| 159 | Triglycerides in IDL | 0.01837999 |
| 160 | Phospholipids to Total Lipids in Chylomicrons and Extremely Large VLDL percentage | 0.01809273 |
| 161 | Free Cholesterol to Total Lipids in Small VLDL percentage | 0.0174741 |
| 162 | Triglycerides to Total Lipids in Medium VLDL percentage | 0.01733313 |
| 163 | Triglycerides in Medium VLDL | 0.01720896 |

| | | |
|---|---|---|
| 164 | Concentration of Large LDL Particles | 0.01717995 |
| 165 | Free Cholesterol in Small HDL | 0.01702882 |
| 166 | Omega-3 Fatty Acids | 0.01698015 |
| 167 | Triglycerides in HDL | 0.01694551 |
| 168 | Average Diameter for VLDL Particles | 0.01681538 |
| 169 | Phospholipids to Total Lipids in Very Large HDL percentage | 0.01645793 |
| 170 | Triglycerides to Total Lipids in Large LDL percentage | 0.01625255 |
| 171 | Concentration of Medium LDL Particles | 0.01551486 |
| 172 | Free Cholesterol in HDL | 0.0154946 |
| 173 | Cholesterol to Total Lipids in Small HDL percentage | 0.01538546 |
| 174 | Monounsaturated Fatty Acids | 0.0153794 |
| 175 | Free Cholesterol in IDL | 0.01527611 |
| 176 | Triglycerides to Total Lipids in IDL percentage | 0.01504465 |
| 177 | Cholesteryl Esters in Large VLDL | 0.0146185 |
| 178 | Triglycerides to Total Lipids in Large VLDL percentage | 0.01458102 |
| 179 | Triglycerides to Total Lipids in Small LDL percentage | 0.01390909 |
| 180 | Free Cholesterol in Medium HDL | 0.01376349 |
| 181 | Free Cholesterol to Total Lipids in Medium HDL percentage | 0.01372066 |
| 182 | Saturated Fatty Acids | 0.01311657 |
| 183 | Triglycerides in Small VLDL | 0.0130379 |
| 184 | Basophill count | 0.01291488 |
| 185 | Cholesteryl Esters to Total Lipids in Medium VLDL percentage | 0.012909 |
| 186 | High light scatter reticulocyte count | 0.01248926 |
| 187 | Total Lipids in IDL | 0.01234108 |
| 188 | Triglycerides to Total Lipids in Medium LDL percentage | 0.01232081 |
| 189 | Phospholipids in Small LDL | 0.0122887 |
| 190 | Triglycerides to Total Lipids in Very Large HDL percentage | 0.01222629 |
| 191 | Triglycerides to Total Lipids in Small VLDL percentage | 0.01221344 |
| 192 | Triglycerides to Total Lipids in Chylomicrons and Extremely Large VLDL percentage | 0.01199153 |

| | | |
|---|---|---|
| 193 | Total Phospholipids in Lipoprotein Particles | 0.01178749 |
| 194 | LDL direct | 0.01155847 |
| 195 | Cholesterol in Small VLDL | 0.01131319 |
| 196 | Concentration of IDL Particles | 0.01110491 |
| 197 | Average Diameter for HDL Particles | 0.01105896 |
| 198 | Remnant Cholesterol (Non-HDL, Non-LDL -Cholesterol) | 0.01104967 |
| 199 | Free Cholesterol to Total Lipids in Medium VLDL percentage | 0.01095202 |
| 200 | Total Fatty Acids | 0.01093665 |
| 201 | Cholesteryl Esters in Medium VLDL | 0.01093092 |
| 202 | Cholesteryl Esters in Large HDL | 0.01062089 |
| 203 | Cholesteryl Esters to Total Lipids in Large VLDL percentage | 0.01037524 |
| 204 | Phospholipids in Very Large HDL | 0.01035061 |
| 205 | Cholesterol to Total Lipids in Very Large VLDL percentage | 0.01027581 |
| 206 | Cholesteryl Esters in Very Large VLDL | 0.01017354 |
| 207 | Cholesteryl Esters in Chylomicrons and Extremely Large VLDL | 0.01013106 |
| 208 | Triglycerides in Very Small VLDL | 0.00987373 |
| 209 | Triglycerides in Large VLDL | 0.00955688 |
| 210 | Free Cholesterol to Total Lipids in Small LDL percentage | 0.00945337 |
| 211 | Cholesterol to Total Lipids in Large VLDL percentage | 0.00941908 |
| 212 | Cholesteryl Esters in IDL | 0.00916918 |
| 213 | Triglycerides in Small HDL | 0.00915968 |
| 214 | Triglycerides in Medium LDL | 0.00903231 |
| 215 | Cholesteryl Esters to Total Lipids in Small HDL percentage | 0.00881831 |
| 216 | Free Cholesterol in Small LDL | 0.00880436 |
| 217 | Free Cholesterol in Chylomicrons and Extremely Large VLDL | 0.00867329 |
| 218 | Free Cholesterol to Total Lipids in Large LDL percentage | 0.00861364 |
| 219 | Phospholipids to Total Lipids in Small VLDL percentage | 0.00852171 |
| 220 | Total Lipids in Small VLDL | 0.00848308 |
| 221 | Triglycerides to Total Lipids in Very Small VLDL percentage | 0.00835423 |

| | | |
|---|---|---|
| 222 | Cholesteryl Esters in Very Small VLDL | 0.0082757 |
| 223 | Cholesteryl Esters to Total Lipids in Very Small VLDL percentage | 0.00821849 |
| 224 | Cholesterol in Large HDL | 0.00818894 |
| 225 | Triglycerides in LDL | 0.00815025 |
| 226 | Phospholipids in Medium LDL | 0.00812496 |
| 227 | Cholesterol in Very Large HDL | 0.00798942 |
| 228 | Free Cholesterol in Medium LDL | 0.00795098 |
| 229 | Phospholipids in Very Small VLDL | 0.00787569 |
| 230 | Cholesterol in Very Small VLDL | 0.00782841 |
| 231 | Cholesterol in Large LDL | 0.00781733 |
| 232 | Cholesteryl Esters in Very Large HDL | 0.00755785 |
| 233 | Cholesterol in Medium VLDL | 0.00746452 |
| 234 | Triglycerides in Chylomicrons and Extremely Large VLDL | 0.0073954 |
| 235 | Cholesterol in IDL | 0.00738456 |
| 236 | Free Cholesterol in Small VLDL | 0.00717628 |
| 237 | Phospholipids in IDL | 0.007075 |
| 238 | Concentration of Very Small VLDL Particles | 0.00701662 |
| 239 | Free Cholesterol in Very Small VLDL | 0.00701241 |
| 240 | Concentration of Chylomicrons and Extremely Large VLDL Particles | 0.00698035 |
| 241 | Phospholipids in Large HDL | 0.0069734 |
| 242 | Cholesterol to Total Lipids in Medium VLDL percentage | 0.00691197 |
| 243 | Triglycerides to Total Lipids in Very Large VLDL percentage | 0.0068699 |
| 244 | Concentration of Large HDL Particles | 0.00679131 |
| 245 | Phospholipids in Small VLDL | 0.00678364 |
| 246 | Clinical LDL Cholesterol | 0.00677877 |
| 247 | Cholesteryl Esters in Medium LDL | 0.00673466 |
| 248 | Free Cholesterol in Large HDL | 0.00666778 |
| 249 | Cholesteryl Esters in Small VLDL | 0.00637898 |
| 250 | Phosphoglycerides | 0.00629068 |

| | | |
|---|---|---|
| 251 | Total Esterified Cholesterol | 0.00626451 |
| 252 | Cholesterol to Total Lipids in Very Small VLDL percentage | 0.00607715 |
| 253 | VLDL Cholesterol | 0.00595139 |
| 254 | Total Cholines | 0.00568314 |
| 255 | Phospholipids in Medium VLDL | 0.00568289 |
| 256 | Total Lipids in Small LDL | 0.00538762 |
| 257 | Free Cholesterol in VLDL | 0.00530808 |
| 258 | Total Lipids in Very Small VLDL | 0.00523371 |
| 259 | Cholesterol in Large VLDL | 0.00516661 |
| 260 | Cholesteryl Esters in VLDL | 0.00512531 |
| 261 | Total Lipids in Lipoprotein Particles | 0.00504153 |
| 262 | Cholesterol in Very Large VLDL | 0.00501382 |
| 263 | Total Lipids in Large HDL | 0.00491935 |
| 264 | Phospholipids in Chylomicrons and Extremely Large VLDL | 0.00482207 |
| 265 | Triglycerides in Very Large VLDL | 0.00479994 |
| 266 | Concentration of Medium VLDL Particles | 0.00471031 |
| 267 | Total Lipids in Large VLDL | 0.00470646 |
| 268 | Free Cholesterol in Very Large VLDL | 0.00468838 |
| 269 | Total Lipids in Chylomicrons and Extremely Large VLDL | 0.00460196 |
| 270 | Free Cholesterol in LDL | 0.004392 |
| 271 | Triglycerides in Small LDL | 0.00438247 |
| 272 | Total Free Cholesterol | 0.00436302 |
| 273 | Phospholipids in VLDL | 0.00434042 |
| 274 | Free Cholesterol in Large VLDL | 0.00431186 |
| 275 | Triglycerides in VLDL | 0.00427312 |
| 276 | Cholesterol in Medium LDL | 0.00425774 |
| 277 | Free Cholesterol in Large LDL | 0.00419364 |
| 278 | Total Lipids in Very Large VLDL | 0.00416661 |
| 279 | Total Cholesterol | 0.00414861 |

| | | |
|---|---|---|
| 280 | Apolipoprotein B | 0.00412019 |
| 281 | Cholesterol in Small LDL | 0.00409213 |
| 282 | Free Cholesterol in Medium VLDL | 0.00403143 |
| 283 | Total Lipids in VLDL | 0.00392858 |
| 284 | Cholesteryl Esters in Small LDL | 0.00389343 |
| 285 | Phospholipids in Large VLDL | 0.00371735 |
| 286 | Total Lipids in Medium LDL | 0.00341809 |
| 287 | Concentration of Large VLDL Particles | 0.00340988 |
| 288 | Concentration of VLDL Particles | 0.00336871 |
| 289 | LDL Cholesterol | 0.00335124 |
| 290 | Cholesterol in Chylomicrons and Extremely Large VLDL | 0.00334478 |
| 291 | Total Cholesterol Minus HDL-C | 0.00327428 |
| 292 | Total Triglycerides | 0.00321507 |
| 293 | Concentration of LDL Particles | 0.00320291 |
| 294 | Cholesteryl Esters in LDL | 0.00319469 |
| 295 | Phospholipids in LDL | 0.0031911 |
| 296 | Phospholipids in Large LDL | 0.00283319 |
| 297 | Cholesteryl Esters in Large LDL | 0.00258274 |
| 298 | Total Lipids in Large LDL | 0.00243433 |
| 299 | Phospholipids in Very Large VLDL | 0.00216243 |
| 300 | Concentration of Very Large VLDL Particles | 0.0021317 |
| 301 | Total Lipids in LDL | 0.00201787 |
| 302 | Nucleated red blood cell count | 0.0004003 |
| 303 | Nucleated red blood cell percentage | 0.00011636 |

16. **Supplementary Figure F1:** A histogram of alcohol consumption in terms of DPW across sex. As shown males dominate the heavy drinking spectrum compared to females.

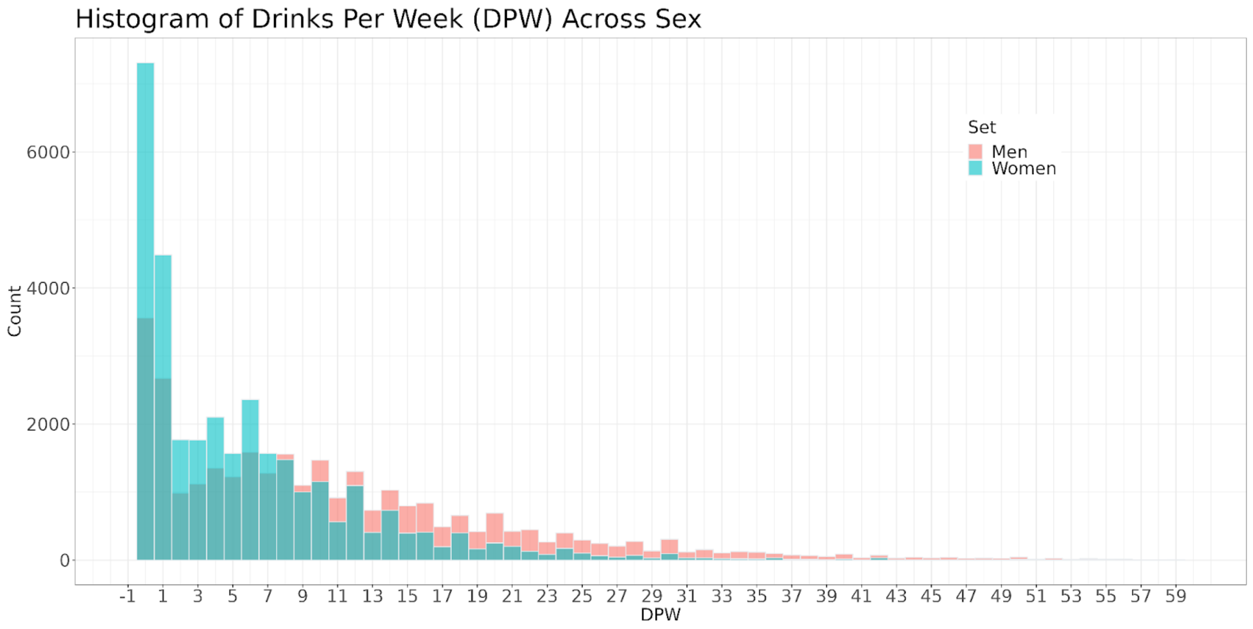

17. **Supplementary Figure F2:** A dendrogram plot of the full list of the blood biomarkers used to predict DPW. Variables that are close to each other represent a strong relationship.

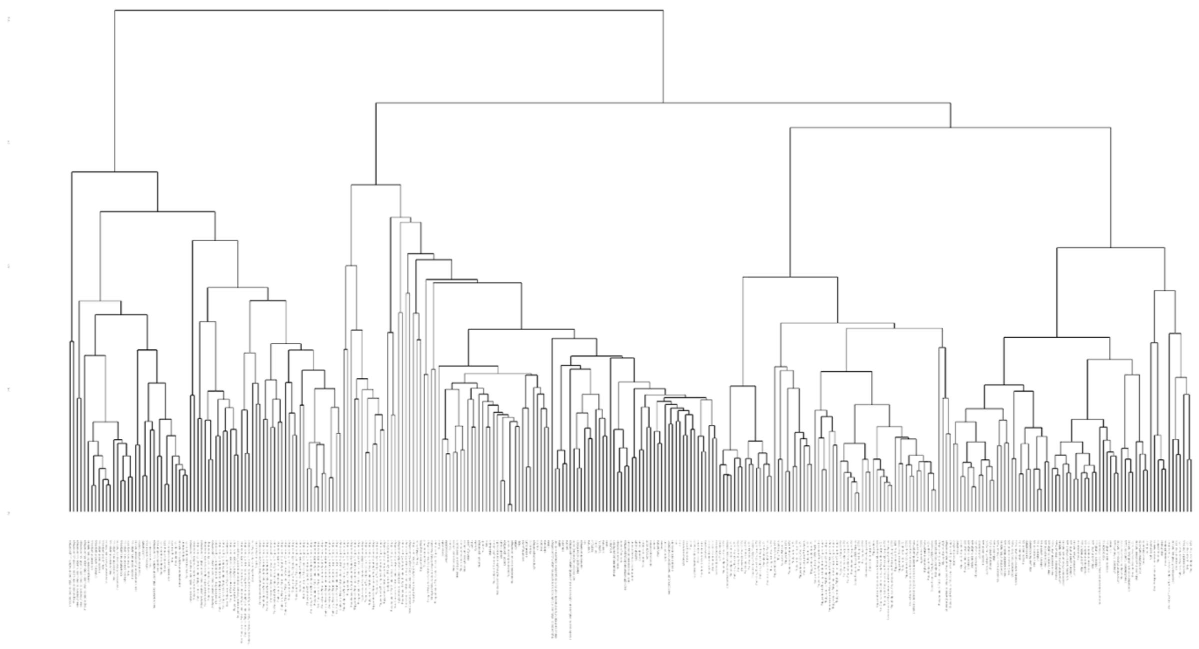
